# External validation of prognostic and predictive gene signatures in head and neck cancer patients

**DOI:** 10.1101/2024.09.24.24314278

**Authors:** Erlend I. F. Fossen, Mauricio Moreira-Soares, Marissa LeBlanc, Arnoldo Frigessi, Eivind Hovig, Laura Lopez-Perez, Estefanía Estévez-Priego, Liss Hernandez, Maria Fernanda Cabrera-Umpierrez, Giuseppe Fico, Ingeborg Tinhofer, Vanessa Sachse, Kathrin Scheckenbach, Christophe Le Tourneau, Maud Kamal, Steve Thomas, Miranda Pring, Lisa Licitra, Loris De Cecco, Stefano Cavalieri

**Affiliations:** Oslo Centre for Biostatistics and Epidemiology, Institute of Basic Medical Sciences, University of Oslo, Oslo, Norway; CICERO Center for International Climate Research, Oslo, Norway; Department of Method Development and Analytics, Norwegian Institute of Public Health, Oslo, Norway; Centre for Bioinformatics, Department of Informatics, University of Oslo, Oslo, Norway; Department of Tumor biology, Institute for Cancer Research, Oslo University Hospital, 0310 Oslo, Norway; Universidad Politécnica de Madrid-Life Supporting Technologies Research Group, ETSIT, 28040 Madrid, Spain; Department of Radiooncology and Radiotherapy, Translational Radiation Oncology Research Laboratory, Charité – Universitätsmedizin Berlin, corporate member of Freie Universität Berlin and Humboldt Universität zu Berlin, Charitéplatz 1, 10117 Berlin, Germany; Heinrich-Heine-Universität Düsseldorf, Dusseldorf, Germany; Department of Drug Development and Innovation (D3i), Institut Curie, Paris-Saclay University, Paris, France; Bristol Dental School, University of Bristol, United Kingdom; Head and Neck Medical Oncology Department, Fondazione IRCCS Istituto Nazionale dei Tumori di Milano, via Giacomo Venezian 1, 20133 Milan, Italy; Department of Oncology and Hemato-oncology, University of Milan, via Santa Sofia 9/1, 20122 Milan, Italy; Experimental Oncology, Fondazione IRCCS Istituto Nazionale dei Tumori, via Giacomo Venezian 1, 20133 Milan, Italy

## Abstract

Head and neck squamous cell carcinomas (HNSCC) are aggressive and heterogenous tumors with a high fatality rate. Many gene signatures (GS) have been developed with both prognostic and predictive significance. We aimed to externally validate five published GS in a large European collection of HNSCC patients. Gene expression from 1097 treatment-naïve HNSCC patients’ primary tumors was used to calculate scores for the five GS. Cox proportional hazard models were used to test the association between both 2-year overall survival and 2-year disease-free survival and the signature scores. The predictive role of GS was validated by comparing survival associations in patients receiving specific treatment (i.e., radiotherapy, systemic treatment) versus those who did not. We successfully externally validated all 5 GS, including two prognostic signatures, one signature as prognostic and predictive of sensitivity to systemic treatment, while signatures for cisplatin-sensitivity and radiosensitivity were validated as prognostic only.

## 1. Introduction

Head and neck cancer (HNC) is the 7^th^ most common cancer worldwide, with more than 890,000 new cases and 450,000 deaths annually^1,2^. Head and neck squamous cell carcinoma (HNSCC) is the most common histological type of HNC, making up 90% of all HNC cases^2,3^, roughly 4.5% of all global cancer diagnoses and approximately 4.6% of all cancer deaths^1,2^. Major risk factors for HNSCC are tobacco smoking and alcohol consumption^1,3^, but human papillomavirus (HPV) infection is also a known risk factor for oropharyngeal cancers^4^.

HNSCCs are a heterogenous group of cancers with malignancies developing at various anatomical sub-sites and with high molecular tumor heterogeneity^5,6^. This heterogeneity contributes to differences between patients in terms of treatment response with a need for personalized approaches^6,7^. In general, HNSCC patients with early disease (stage I-II) have high cure rates with single modality therapies (either surgery or radiotherapy). Subjects with loco-regionally advanced disease (stage III-IVa/b) are treated with multimodal approaches (i.e., various combinations of surgery, radiotherapy and systemic treatment)^8^. Despite optimal treatments, approximately half of patients develop recurrence within 2 years and half die with disease within 5 years from the diagnosis^9^. Reliable biomarkers and gene signatures (GS) are crucial for developing personalized treatment^10,11^. GS are sets of genes involved in a biological process that can provide information about the expected disease outcome (prognostic) and/or the response to a specific treatment (predictive)^10,11^. Prognostic signatures provide information about disease prognosis irrespective of therapy. Meanwhile, predictive signatures inform about how likely a patient is to respond to a specific therapy and can therefore enable more personalized treatment plans.

Many prognostic GS, calculated from gene expression in treatment-naïve tumors, have been developed for HNSCC^12–16^. External validation of already published GS is an important step towards incorporating GS in clinical practice^17^. In this study, we decided to consider only GS which were already published, and which were either disease- or treatment-specific. Among them is a 172-gene signature^18^ (*172-GS*) that was developed to be prognostic of patient’s risk of relapse in HNSCC patients independently of HPV-status, but was later shown to not be prognostic of overall survival (OS) in HPV-positive patients^19^. However, in the same patient population, a specific three-cluster HPV signature^20^ (*3 clusters HPV*) was developed and externally validated as prognostic of OS^19^. As for predictive signatures, the 10-gene radiosensitivity index^21^ (*RSI*) is among the most validated pan-cancer GSs for prediction of sensitivity to radiotherapy^22^. Another pan-cancer predictive signature, in this case predictive of cisplatin-sensitivity^23^ (*pancancer-cisplatin*), was recently developed, but has not been externally validated in HNSCC patients. Lastly, a signature that classifies HNC patients into six different subtypes/clusters was developed by De Cecco et al^24^. One of them (*Cl3-hypoxia*) showed hypoxic features and was found to be prognostic^19,25^ and related to response to treatment with anti-EGFR agents (cetuximab and afatinib)^26,27^.

Using a large collection of HNSCC patients with available gene expression data, we aimed to externally validate two prognostic gene signatures (*172-GS* and *3 clusters HPV*), two predictive gene signatures (*RSI* and *pancancer-cisplatin*) and one prognostic and predictive GS (*Cl3-hypoxia*). In contrast to earlier studies, we wanted to explicitly test whether the prognostic signatures (*172-GS* and *3 clusters HPV*) are valid independent of HPV-status, and explicitly test if the predictive signatures are predictive of treatment response by comparing if the signature effect is only found in patients with a specific treatment and not in patients without that treatment. We tested these signatures using two survival endpoints, OS and disease-free survival (DFS).

## 2. Methods

### 2.1. Ethics statement

This study was conducted in full accordance with the World Medical Association’s Declaration of Helsinki (2013 version). The protocol was ethically approved by the Norwegian Regional Committee for Medical and Health Research Ethics (REK) South-East A under application number 270467. The data is securely stored in the University of Oslo’s server for sensitive data (TSD/USIT), adhering to the requirements of GDPR legislation.

Access to the data is granted only to authorized collaborators who have been included in the ethical approval. All proprietary studies that contributed data obtained ethical approval from their respective local authorities in Italy, Germany, or France. Copies of these ethical approvals were provided to the principal investigator by the data providers. BD2Decide was approved by the Ethical Committee of the Fondazione IRCCS Istituto Nazionale dei Tumori (Milan, Italy) in 2016 and it has two identifiers: INT65-16 and INT66-16. Biomarker analysis of the ARO 04-01 Def-RCT cohort was approved by the Ethical Committee of the Charité - Universitätsmedizin Berlin (Berlin, Germany) in 2010 (EA2/086/10). Biomarker analysis of the DKTKRO Def-RCT was approved by the Ethical Committee of the Technical University of Dresden (Dresden, Germany) in 2014 (EK200112014).

### 2.2. Patient selection

Within a European cooperative research project named SuPerTreat (research grant nr. ERAPERMED2019-281, further details in Funding), we first identified and constructed a multicenter dataset, which consisted primarily of data collected in previous research projects^28–36^ (NCT02832102, NCT03017573, NCT02059668) at the Fondazione IRCCS Istituto Nazionale dei Tumori (Milan, Italy), Institut Curie (Paris, France) and Charité - Universitätsmedizin Berlin (Berlin, Germany) (Table S1 in supplementary information). We additionally included two public datasets (TCGA^37,38^ and GSE41613^39^) that adhered to MIAME^40^ and MINSEQE^41^ standards.

The selected studies complied with the following **inclusion criteria**:

1. Diagnosis of head and neck squamous cell carcinoma (HNSCC)
2. Planned treatment with curative intent
3. Tumor specimens (formalin-fixed paraffin-embedded, FFPE) of HNSCC sampled prior to treatment, already assessed for gene expression analysis
4. Stage I, II, III, IVA or IVB according to the tumor-node-metastasis (TNM) cancer staging system 7th edition^42^
5. One of the following tumor sites: oral cavity, oropharynx, hypopharynx, larynx
6. Age ≥18 years

A total of N = 1589 patients were screened from these studies. From these, patients were excluded based on the following **general exclusion criteria**:

1. Gene expression data not available
2. Distant metastasis at diagnosis or missing information about stage
3. Recurrent/metastatic disease at the time of study entry
4. Missing information about survival timing or survival status

**Additional exclusion criteria** were applied when testing gene signatures related to sensitivity to systemic treatment:

5. Early disease stage excluded when testing signatures *pancancer-cisplatin* and *Cl3-hypoxia*. This specification is due to the fact that most early disease stage HNSCC patients are not treated with systemic treatment
6. Missing treatment information about systemic treatment agent when testing signatures *pancancer-cisplatin* and *Cl3-hypoxia*

A total of 1097 patients were eligible for inclusion in this study, with the number reduced for gene signatures related to sensitivity to systemic treatment (Figure 1, Table S1). In models testing the signatures *172-GS*, *3 clusters HPV* and *RSI*, all 1097 patients were eligible for analyzing OS (Figure 1) and 907 patients were eligible for DFS (Figure S1). In models testing sensitivity to systemic treatment signatures (*pancancer-cisplatin* and *Cl3-hypoxia*), 750 patients were eligible for analyses of OS (Figure S2) and 700 patients were eligible for analyses of DFS (Figure S3).

**Figure 1.**
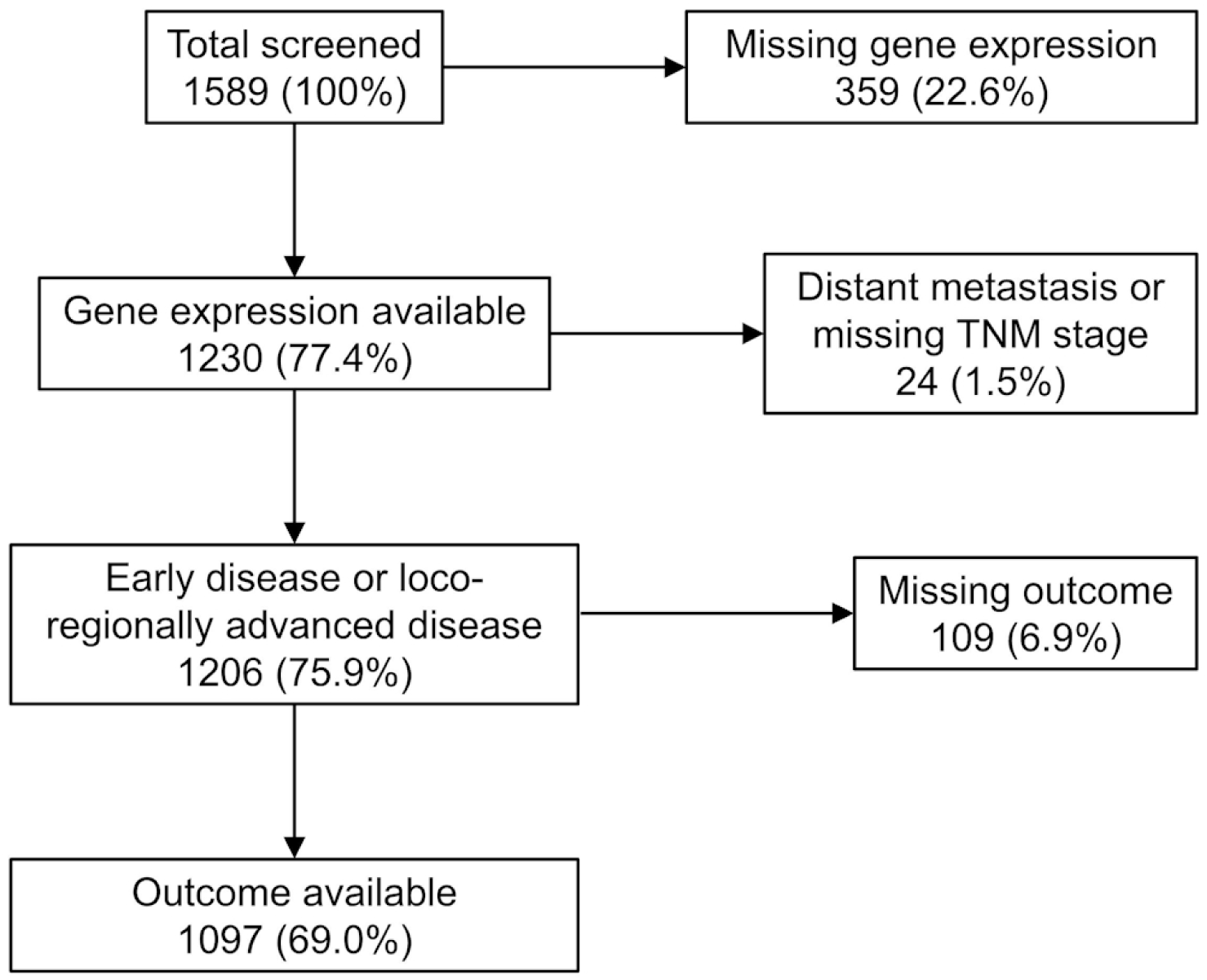
Exclusion flowchart. The largest number of eligible patients used in the most inclusive models (models testing the signatures *172-GS*, *3 clusters HPV* and *RSI* with overall survival as the endpoint) is shown. See Figures S1, S2, S3 for exclusion flowcharts for other gene signature models.

None of the patients that were eligible for testing the signatures *172-GS*, *RSI*, *pancancer-cisplatin* or *Cl3-hypoxia* were previously included in the development of these signatures. For the *3 clusters HPV* GS, most of the eligible HPV-positive patients (N = 152, 14% of all eligible patients) were already part of a previous external validation of the signature^19^, but in this study we additionally tested the GS in HPV-negative patients and adjusted for more covariates.

### 2.3. Endpoints, clinical variables and data harmonization

OS, defined as survival time from diagnosis until death of any cause or censoring, was recorded in all studies. DFS, defined as time from diagnosis until cancer recurrence or death of any cause, whichever occurred first, or censoring, was recorded in all but two studies (Table S1). Patients were followed for up to 20 years in the period 1989-2017. Co-primary endpoints were 2-year OS and 2-year DFS.

GSs were considered prognostic if they were associated with general disease prognosis independently of treatments. We defined a GS as predictive if the prognostic capability was limited to a population receiving a specific treatment.

Data harmonization of clinical variables was performed following head and neck ontology (HeNeCOn^43^) and quality rules defined in BD2Decide study^28^ (extended in the context of the SuPerTreat project), to achieve comparable measurements across studies (see Table 1 and Tables S2 and S3 for full list of clinical covariates). Tumor staging at diagnosis was performed according to the 7th edition of the AJCC/UICC staging system^42^. The staging variable was then classified in two main groups based on disease extension: early disease (stage I-II) vs. loco-regionally advanced disease (stage III-IVa/b). Human papillomavirus (HPV) infection is a known risk factor for oropharyngeal cancer^4^ and HPV-status was recorded for tumor tissues of all patients with oropharyngeal cancer, using either HPV DNA/RNA tests (with DNA PCR or ISH with or without E6/E7 mRNA confirmation, as per local guidelines) or through p16ink4a immunostaining. Non-oropharyngeal HNSCCs (including oral cavity cancers^44^) were considered HPV-negative in analyses. Systemic treatments were coded in two ways: 1) platinum-based (if carboplatin and/or cisplatin were part of the agents a patient received) or non-platinum based (primarily cetuximab or paclitaxel); and 2) cetuximab-based (if cetuximab was part of the agents a patient received) or non-cetuximab based. Patients not receiving systemic treatments were in both cases coded as such.

**Table 1.** Cohort characteristics for head and neck cancer patients used to test the *172-GS* and *3 clusters HPV* signatures. The disease-free survival (DFS) dataset is a subset of the overall survival dataset, where patients without data on DFS are excluded.

|  | Overall survival dataset<br>(N=1097) | Disease-free survival dataset<br>(N=907) |
| --- | --- | --- |
| **172-GS score** |  |  |
| Mean (SD) | 0.0 (1.0) | 0.0 (1.0) |
| Median (Min, Max) | 0.1 (-4.5, 3.6) | 0.1 (-4.9, 3.5) |
| **3 clusters HPV score** |  |  |
| Mean (SD) | 0.0 (1.0) | 0.0 (1.0) |
| Median (Min, Max) | 0.0 (-2.8, 6.9) | 0.0 (-2.9, 3.5) |
| **Study ID** |  |  |
| BD2_INT_MI | 310 (28.3%) | 310 (34.2%) |
| BD2_UDUS | 127 (11.6%) | 127 (14.0%) |
| GECO110 | 107 (9.8%) | 107 (11.8%) |
| GSE41613 | 97 (8.8%) | 0 (0%) |
| SCANDARE | 32 (2.9%) | 0 (0%) |
| TCGA | 424 (38.7%) | 363 (40.0%) |
| **Clinical age at diagnosis** |  |  |
| Mean (SD) | 61.0 (11.9) | 60.8 (11.7) |
| Median (Min, Max) | 61.0 (19.0, 93.0) | 61.0 (19.0, 93.0) |
| **Clinical sex** |  |  |
| Male | 794 (72.4%) | 660 (72.8%) |
| Female | 303 (27.6%) | 247 (27.2%) |
| **Disease extension at diagnosis** |  |  |
| Early disease | 161 (14.7%) | 98 (10.8%) |
| Locoregionally advanced disease | 936 (85.3%) | 809 (89.2%) |
| **Tumor region** |  |  |
| Oropharynx | 227 (20.7%) | 218 (24.0%) |
| Hypopharynx | 55 (5.0%) | 52 (5.7%) |
| Larynx | 165 (15.0%) | 150 (16.5%) |
| Oral cavity | 598 (54.5%) | 451 (49.7%) |
| Missing | 52 (4.7%) | 36 (4.0%) |
| **HPV status** |  |  |
| Positive | 156 (14.2%) | 151 (16.6%) |
| Negative | 941 (85.8%) | 756 (83.4%) |
| **Smoking** |  |  |
| Never | 230 (21.0%) | 213 (23.5%) |
| Current or Former | 653 (59.5%) | 609 (67.1%) |
| Missing | 214 (19.5%) | 85 (9.4%) |
| **Undergone cancer surgery** |  |  |
| No | 187 (17.0%) | 187 (20.6%) |
| Yes | 854 (77.8%) | 672 (74.1%) |
| Missing | 56 (5.1%) | 48 (5.3%) |
| **Radiotherapy treatment** |  |  |
| No | 208 (19.0%) | 146 (16.1%) |
| Yes | 752 (68.6%) | 633 (69.8%) |
| Missing | 137 (12.5%) | 128 (14.1%) |
| **Systemic treatment** |  |  |
| No | 302 (27.5%) | 233 (25.7%) |
| Yes | 461 (42.0%) | 426 (47.0%) |
| Missing | 334 (30.4%) | 248 (27.3%) |

Treatments (surgery, radiotherapy and systemic therapy) performed with curative intent were recorded as received or not received. This is potentially problematic since Cox regression survival models assume that covariates are measured at baseline (diagnosis) or require information about the timing of treatment. The use of the received treatment as a proxy for intended treatment (a baseline covariate) may result in immortal time bias^45^. We performed extensive sensitivity analyses to evaluate if our results were sensitive to using received treatment as a covariate (Appendix 1). First, we considered the degree of mortality observed during the period prior to when treatments were initiated. Second, we used an external dataset (Head and Neck 5000^46,47^) with similar patient characteristics as SuPerTreat and compared treatment coefficients from Cox models with either intended treatment or received treatment as covariates. Third, we compared treatment coefficients from Cox models with received treatment with models where treatment was considered time-dependent (in a subset of patients where the timing of surgery was known). Fourth, we used a landmark analysis^48,49^ and compared treatment coefficients from a Cox model with received treatment with a landmark model where patients without an event or censoring prior to a landmark time point (corresponding with a time at which all patients started treatment) were followed from the landmark time. Lastly, we used quantitative bias analyses^50^ to compare estimates of received treatments with bias-adjusted estimates of treatments. Based on these sensitivity analyses (Results section 3.3.1), we inferred that our models are robust to violations of the assumptions of the Cox regression models, and therefore used received treatment as a covariate in our survival models. Although gene signatures are tested in interaction with treatments, the tumor tissue used to obtain gene signature scores were sampled at diagnosis (prior to treatment).

### 2.4. Bioinformatics analysis

Gene expression data was profiled on different platforms depending on the study. Three studies (study IDs: BD2_INT_MI, BD2_UDUS, GSE41613) were profiled on Affymetrix microarrays (platforms GPL23126 and GPL570), one study (GECO110) was profiled on Illumina microarray (platform GPL14951), and two studies (SCANDARE, TCGA) were profiled using Illumina RNA-seq. Within studies, Affymetrix data were quantile-normalized using robust multi-array average (RMA). Illumina microarray data were quantile-normalized. For RNA-seq data, within studies, we first removed very lowly expressed genes, where genes were removed if all samples had < 0.25 counts per million (cpm), corresponding to max 5 reads in the smallest sample and max 22 reads in the largest sample. RNA-seq data were then quantile-normalized using the limma R package^51^ (function: voom). We used WGCNA package^52^ (function collapseRows and “maxRowVariance” method) to select the most variable probe in cases when multiple probes mapped to the same EntrezID. Probes and ensemble IDs that mapped to multiple EntrezID were removed. All gene expression data were log2-transformed after the abovementioned normalization. Genes that were missing for all patients in a study were removed when combining datasets. ComBat^53,54^ was used to remove batch effects introduced by systemic non-biological technical errors from combining data from multiple studies with different platforms. The largest proprietary study (ID: BD2_INT_MI) was used as reference in ComBat, parametric adjustment was performed, and since there is variability in patient characteristics between studies, we only adjusted for the mean to not remove biological variance between the studies. To avoid bias in downstream analyses^55^, we only included the batches (studies) as covariates in ComBat.

We calculated 5 gene signature scores for each patient following descriptions given in the original publications (see Table S4 for detailed list of genes and weights per signature). The signatures *172-GS*^18^, *3 clusters HPV*^19,20^, *pancancer-cisplatin*^23^, and *Cl3-hypoxia*^24^ were calculated as weighed sums using the equation: 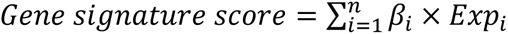, where *βi* is the weight of the i-th gene (of n genes) and *Exp_i_* is the gene expression of the i-th gene. The *172-GS* signature was calculated using the hacksig package^56^. The radiosensitivity index (*RSI*) signature is based on the expression of 10 specific genes. *RSI* was calculated as a weighed ranked sum using the equation: 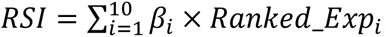, where *βi* is the weight of the *i*-th gene and *Ranked_Exp_i_* is the ranked gene expression of the *i*-th gene. For each patient the 10 genes were ranked in ascending order using patient-specific expression data, where the gene with the lowest expression is assigned a value of 1, and the gene with the highest expression is assigned a value of 10.

Each gene signature was mean-centered and scaled by the standard deviation (SD) within its respective dataset to make the scores easier to interpret (mean = 0, SD = 1, see Appendix 2 for means and SD on original scale). In our OS dataset, 76% of the *172-GS* signature genes were present, 46% of the *3 clusters HPV* signature genes were present, all *RSI* genes were present, 92% of the *pancancer-cisplatin* genes were present, and 94% of the *Cl3-hypoxia* genes were present. For our DFS dataset, the presence of genes was 80% for the *172-GS* signature, 57% for the *3 clusters HPV* signature, all *RSI* genes present, 92% for *pancancer-cisplatin* and 94% for *Cl3-hypoxia* genes (see Appendix 2 for a list of missing genes per gene signature). Any missing genes were missing for all individuals, making regression-based imputation not a viable option. Instead, all missing genes were imputed as having zero expression. Note that the scaled signature values do not change if we instead were to impute with another constant (e.g., imputing the overall mean expression).

### 2.5. Statistical analyses

All statistical analyses were performed in R Statistical Software^57^ (v4.1.3).

#### 2.5.1. Handling of missing clinical covariates

There were missing data in several categorical variables: smoking status, tumor stage and received treatments (i.e., surgery, systemic therapy and radiotherapy). The missingness was largely structural, resulting from some variables not being measured in some of the studies, and therefore regression-based imputation of missing data was not a viable option^58^. Instead, we used a missing indicator where missing values were coded as belonging to a “missing” category (see Table 1 for amounts of missingness).

#### 2.5.2. Clinical base models

Clinical base models, i.e., those not including the gene expression data, were built as benchmark models for models which included gene signatures as covariates. Four Cox proportional hazard models were fit with either 2-year OS or 2-year DFS as endpoints. Two models were fit with the same data used to test signatures not related to sensitivity to systemic treatment (*172-GS*, *3 clusters HPV* and *RSI*) and two models were fit with the same data used to test signatures related to sensitivity to platinum-based or cetuximab-based treatments (*pancancer-cisplatin* and *Cl3-hypoxia*, respectively). In all models, age, sex, tumor region and HPV-status, smoking status, undergone surgery (yes/no), received radiotherapy (yes/no), and study ID were used as covariates. In models testing signatures not related to systemic treatment, disease extension (early stage, locoregionally advanced) and received systemic therapy (yes/no) were also used as covariates. In models testing signatures related to systemic treatment, TNM stage and a variable for systemic treatment (platinum-based version when testing *pancancer-cisplatin*; cetuximab-based version when testing *Cl3-hypoxia*) were included as covariates. The concordance index (Harrell’s C-index^59^) was calculated for each model to compare their discriminatory ability with the ability of models that included gene signatures (but see Hartman et al.^60^ for limitations on interpreting the C-index). Additionally, we calculated each models measure of explained variation (R^2^) as defined by Royston^61^ and building on previous work^62,63^. This measure is similar to R^2^ of linear models but was made for models analyzing censored survival data^61^.

#### 2.5.3. Gene signature validation models

To validate prognostic gene signatures (*172-GS* and *3 clusters HPV*), we fit Cox proportional hazard models where we tested if the gene signature was associated with survival endpoints while adjusting for other covariates. For each GS, we used the same endpoints and covariates as in the clinical base models, but also tested for an interaction between HPV-status and the GS. This allowed us to test if the same association between survival and GSs was found in both HPV-positive and in HPV-negative patients.

To validate predictive gene signatures (*RSI*, *pancancer-cisplatin* and *Cl3-hypoxia*), we fit Cox proportional hazard models where we tested if gene signatures were associated with survival endpoints and if there was evidence that gene signatures modified the effect of the treatments (i.e., testing for an interaction between gene signatures and treatments). Finding a significant interaction where a GS is associated with survival in a specific treatment but not in others would imply that the signature is predictive. For validating each GS, we used the same covariates as in the clinical base models, but also included relevant interactions between GS and treatments. An interaction between radiotherapy and *RSI* was included when validating *RSI*. For validation of the *pancancer-cisplatin* signature, we included an interaction between the *pancancer-cisplatin* signature and systemic treatment. Similarly for validation of *Cl3-hypoxia*, we included an interaction between the *Cl3-hypoxia* signature and systemic treatment (see Appendix 3 for formulas for each GS model).

To test the proportional hazard assumption of our Cox models, we performed sensitivity analyses where we fit equivalent models as described above, using 5-year survival (both for OS and DFS) as the endpoint instead of 2-year survival. We additionally estimated the median follow-up time using the reverse Kaplan-Meier method^64^.

## 3. Results

### 3.1. Cohort characteristics

The largest dataset used for overall survival analyses, representing the full SuPerTreat cohort, contained patients from 6 studies, where 67% of patients were from the two largest studies (Table 1). The majority of patients were men (72%) and the cohort had a median age at diagnosis of 61 years (SD = 12). Most patients had loco-regionally advanced disease (85%) and underwent surgery (78%) and/or radiotherapy treatment (69%), with 42% of patients receiving systemic treatment. There was heterogeneity in tumor regions, and the most frequent primary site was oral cavity (55%) (Table 1). Similar cohort characteristics were observed in the dataset used for disease-free survival (Table 1), and in subsets of patients that were used to test the effect of other GSs on survival endpoints (Tables S2, S3). Among patients who received systemic treatment with information available about the therapeutic agent, 87.6% received platinum-based and 8.5% received cetuximab-based therapies (Table S3).

### 3.2. Gene signatures

#### 3.2.1. Prognostic signature: *172-GS*

The *172-GS* signature was significantly associated with 2-year OS in HPV-negative patients, where increasing the signature score by 1 (representing an increase of 1 standard deviation) resulted in 41% higher hazard (HR = 1.41 [CI: 1.25, 1.59], Table 2, Figure 2). No evidence of an interaction between the signature score and HPV-status was found (p = 0.621), indicating that the association between survival and the GS was similar in HPV-positive patients (HR = 1.87 [CI: 0.61, 5.78], Table 2). However, the score was not significantly associated with OS in HPV-positive patients when testing the marginal effect of the signature conditioned on being HPV-positive (Table 2). When fixing the signature score at zero (representing the mean score), HPV-negative patients had a 4.27-fold higher hazard than HPV-positive patients (Table 2). The models C-index was 0.71 (SE = 0.02) and R^2^ = 0.29. The C-index of the corresponding clinical base model (i.e. not including the GS as a covariate) was 0.69 (SE = 0.02) and R^2^ = 0.23.

**Figure 2.**
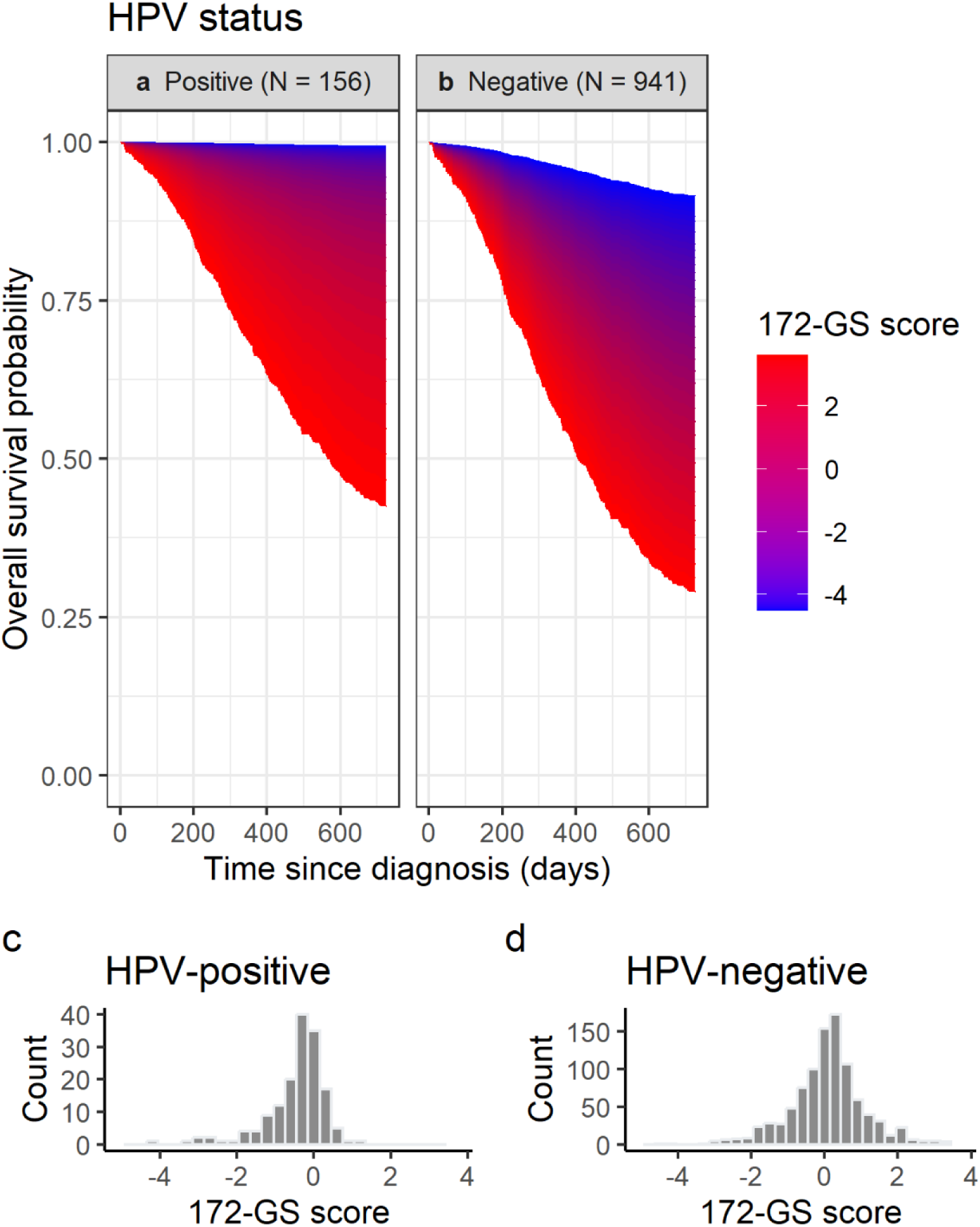
Estimated relationship between the *172-GS* signature and 2-year overall survival. **a, b** Estimated relationship in HPV-positive patients and HPV-negative patients, respectively. The signature score is scaled to a mean of 0 and a standard deviation of 1. Note the difference in sample size (N) per group. **c, d** Distribution of signature values in HPV-positive and HPV-negative patients, respectively.

**Table 2.** Hazard ratios (HR) of gene signatures and covariates that were tested in interaction with the signatures for head and neck cancer patients. Estimates are from models where 2-year overall survival or 2-year disease-free survival was the endpoint and the effect of other clinical covariates were jointly estimated (see Tables S5, S6, S7, S8, S9 for detailed results).

| Signature | Variable | Parameter | Overall survival |  | Disease-free survival |  |
| --- | --- | --- | --- | --- | --- | --- |
|  |  |  | HR [95% CI] | P-value | HR [95% CI] | P-value |
| *172-GS* | HPV-status | HPV positive | Reference | - | Reference | - |
|  |  | HPV negative | 4.27 [2.03, 9.01] | < **0.001** | 4.43 [2.44, 8.02] | < **0.001** |
|  | Signature score | Score given HPV positive | 1.87 [0.61, 5.78] | 0.274 | 1.67 [0.69, 4.05] | 0.254 |
|  |  | Score given HPV negative | 1.41 [1.25, 1.59] | < **0.001** | 1.36 [1.17, 1.59] | < **0.001** |
| *3 clusters HPV* | HPV-status | HPV positive | Reference | - | Reference | - |
|  |  | HPV negative | 4.17 [2.03, 8.57] | < **0.001** | 4.01 [2.26, 7.14] | < **0.001** |
|  | Signature score | Score given HPV positive | 0.41 [0.24, 0.71] | **0.001** | 0.51 [0.33, 0.77] | **0.001** |
|  |  | Score given HPV negative | 0.89 [0.77, 1.02] | 0.084 | 1.06 [0.91, 1.23] | 0.454 |
| *RSI* | Radiotherapy | No | Reference | - | Reference | - |
|  |  | Yes | 0.81 [0.54, 1.21] | 0.306 | 1.08 [0.82, 1.42] | 0.601 |
|  | Signature score | Score given radiotherapy = no | 1.17 [0.90, 1.52] | 0.249 | 1.01 [0.79, 1.27] | 0.929 |
|  |  | Score given radiotherapy = yes | 1.25 [1.08, 1.44] | **0.003** | 1.08 [0.82, 1.42] | 0.306 |
| *Pancancer-cisplatin* | Systemic therapy | Platinum based | Reference | - | Reference | - |
|  |  | Non-platinum based | 0.57 [0.28, 1.15] | 0.114 | 0.94 [0.44, 2.01] | 0.873 |
|  |  | No systemic treatment | 1.24 [0.75, 2.04] | 0.409 | 0.97 [0.63, 1.49] | 0.882 |
|  | Signature score | Score given systemic therapy = platinum based | 1.29 [1.04, 1.59] | **0.020** | 1.23 [0.82, 1.84] | 0.316 |
|  |  | Score given systemic therapy = non-platinum based | 0.82 [0.42, 1.59] | 0.552 | 2.00 [0.90, 4.45] | 0.089 |
|  |  | Score given systemic therapy = no | 1.37 [0.51, 3.69] | 0.540 | 0.92 [0.37, 2.31] | 0.862 |
| *CI3-hypoxia* | Systemic therapy | Cetuximab based | Reference | - | Reference | - |
|  |  | Non-cetuximab based | 1.07 [0.49, 2.31] | 0.869 | 0.71 [0.34, 1.48] | 0.363 |
|  |  | No systemic treatment | 1.22 [0.51, 2.91] | 0.657 | 0.69 [0.31, 1.54] | 0.364 |
|  | Signature score | Score given systemic therapy = cetuximab based | 0.67 [0.39, 1.14] | 0.137 | 0.61 [0.33, 1.13] | 0.114 |
|  |  | Score given systemic therapy = non-cetuximab based | 0.69 [0.54, 0.87] | **0.002** | 0.59 [0.44, 0.80] | **0.001** |
|  |  | Score given systemic therapy = no | 1.70 [1.12, 2.58] | **0.013** | 1.71 [1.14, 2.56] | **0.010** |

Similar effect sizes and significance levels were found for 2-year DFS, with a significant association between the signature score and survival in HPV-negative patients (HR = 1.36 [CI: 1.17, 1.59], Table 2). There was no evidence of an interaction between the signature score and HPV-status (p = 0.651), and the score was not significantly associated with DFS in HPV-positive patients when conditioned on being HPV-positive (Table 2). The models C-index was 0.67 (SE = 0.02) and R^2^ = 0.22. The corresponding clinical base model had a C-index of 0.66 (SE = 0.02) and R^2^ = 0.18. See Table S5 for details about other covariates included in the *172-GS* models.

#### 3.2.2. Prognostic signature: *3 clusters HPV*

There was a significant association between the *3 clusters HPV* signature and 2-year OS in HPV-positive patients, where increasing the signature score resulted in a lower hazard (HR = 0.41 [CI: 0.24, 0.71], Table 2, Figure 3). There was evidence of an interaction between the signature score and HPV-status (p = 0.007), indicating that the effect of the GS was different in HPV-negative patients (HR = 0.89 [CI: 0.77, 1.02], Table 2). Moreover, the score was not significantly associated with OS in HPV-negative patients (Table 2). These patients had a 4.17-fold higher hazard than HPV-positive patients when fixing the signature score at zero (Table 2). The model had a R^2^ = 0.25 and C-index = 0.69 (SE = 0.02). The C-index of the corresponding clinical base model was 0.69 (SE = 0.02) and R^2^ = 0.23.

**Figure 3.**
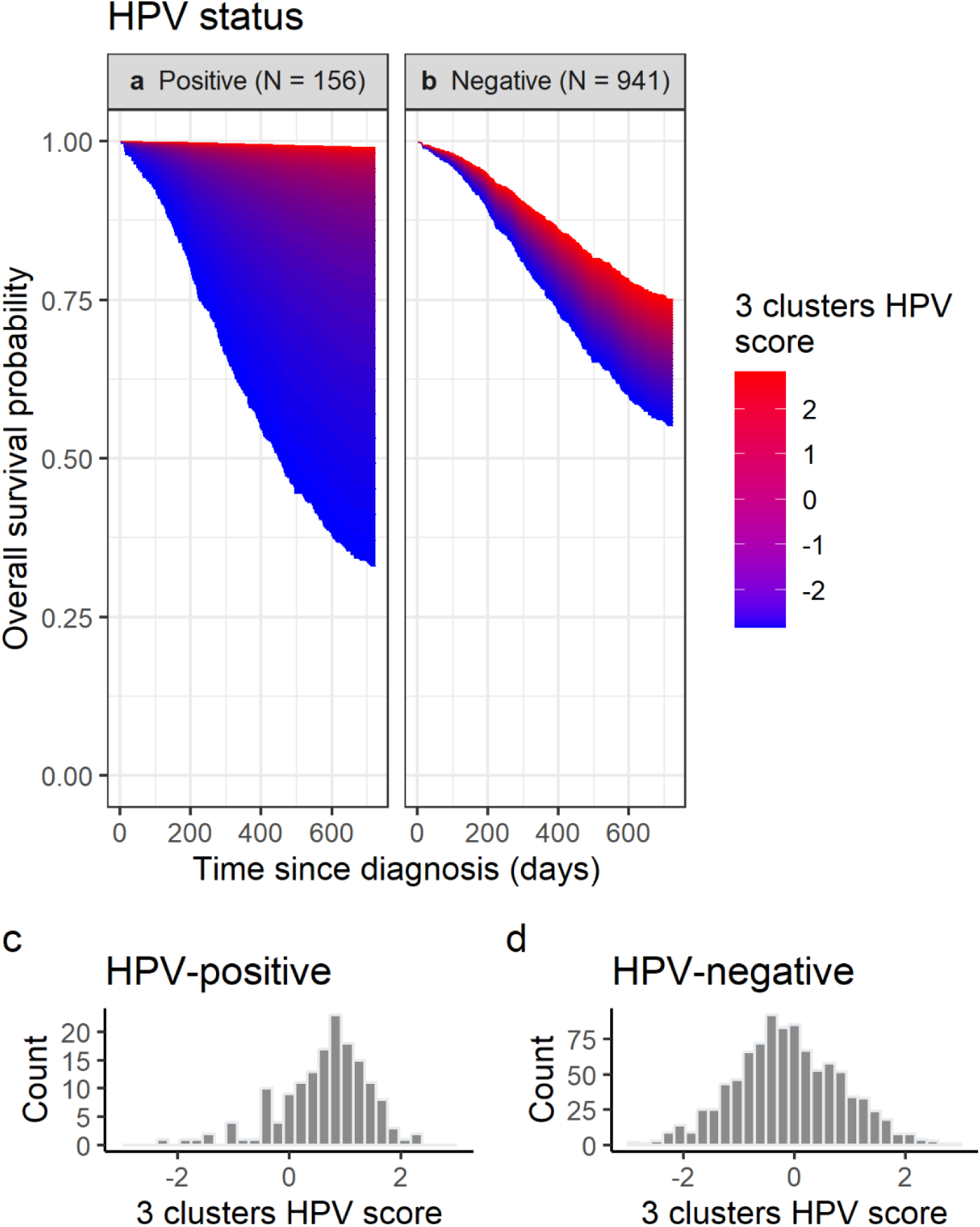
Estimated relationship between the *3 clusters HPV* signature and 2-year overall survival. **a, b** Estimated relationship in HPV-positive patients and HPV-negative patients, respectively. The signature score is scaled to a mean of 0 and a standard deviation of 1. Note the difference in sample size (N) per group. **c, d** Distribution of signature values in HPV-positive and HPV-negative patients, respectively.

We found similar effect sizes and significance levels for 2-year DFS, with a significant association between the GS and survival in HPV-positive patients (HR = 0.51 [CI: 0.33, 0.77], Table 2). Evidence of an interaction between the signature score and HPV-status was also found (p = 0.001), and the score was not significantly associated with DFS in HPV-negative patients (Table 2). The models C-index was 0.67 (SE = 0.02) and R^2^ = 0.20. The C-index of the corresponding clinical base model was 0.66 (SE = 0.02) and R^2^ = 0.18. See Table S6 for details about other covariates included in the *3 clusters HPV* models.

#### 3.2.3. Predictive signature: radiosensitivity index (*RSI*)

The *RSI* was significantly associated with 2-year OS in patients receiving radiotherapy, where a higher hazard was found when increasing the signature score (HR = 1.25 [CI: 1.08, 1.44], Table 2, Figure 4). We found no evidence of an interaction between *RSI* and radiotherapy (p = 0.672), suggesting that the association between survival and *RSI* was the same, although not significantly associated, in patients that did not receive radiotherapy (HR = 1.17 [0.90, 1.52], Table 2, Figure 4). When *RSI* was set to zero, there was no significant association between survival and having received radiotherapy (Table 2). The models C-index was 0.70 (SE = 0.02) and R^2^ = 0.25. The corresponding clinical base model had a C-index of 0.69 (SE = 0.02) and R^2^ = 0.23.

**Figure 4.**
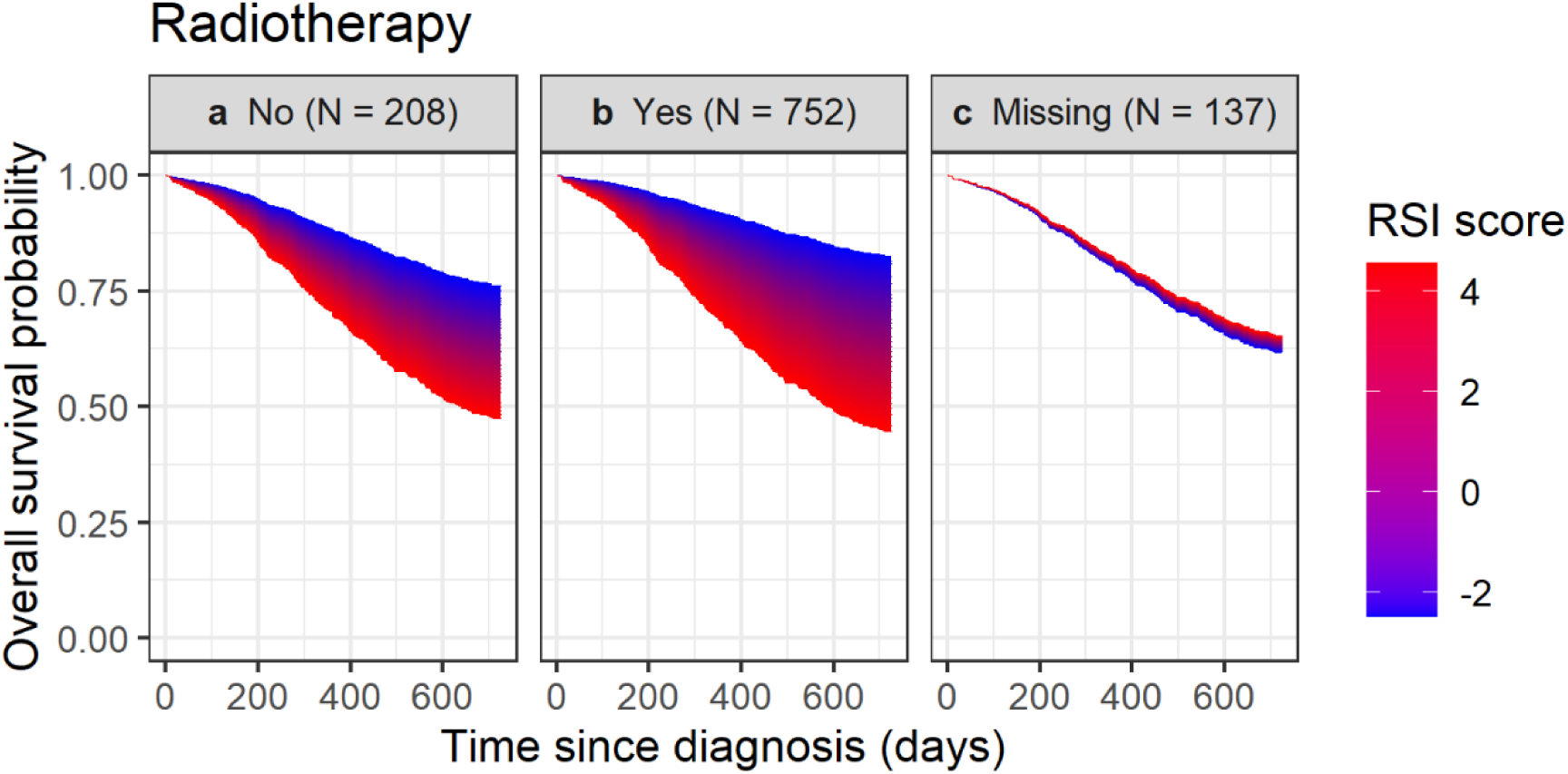
Estimated relationship between the radiosensitivity index (*RSI*) and 2-year overall survival. **a** Relationship in patients that did not receive radiotherapy. **b** Relationship in patients that received radiotherapy. **c** Relationship in patients with missing information about radiotherapy treatment. The *RSI* score is scaled to a mean of 0 and a standard deviation of 1. Note the difference in sample size (N) per group.

In contrast to OS, there was no significant association between 2-year DFS and *RSI* (Table 2). There was also no evidence of an interaction between *RSI* and having received radiotherapy (p = 0.601). The models C-index was 0.66 (SE = 0.02) and R^2^ = 0.18. The corresponding clinical base models C-index was 0.66 (SE = 0.02) and R^2^ = 0.18. See Table S7 for details about other covariates included in the *RSI* models.

#### 3.2.4. Predictive signature: *Pancancer-cisplatin*

There was a significant association between the *pancancer-cisplatin* signature and 2-year OS in patients receiving platinum-based chemotherapy, where a higher hazard was observed when increasing the signature (HR = 1.29 [CI: 1.04, 1.59], Table 2, Figure 5). When platinum-based chemotherapy was the reference, there was no evidence of interactions between the signature and systemic treatment (p = 0.200 when compared to non-platinum based, p = 0.908 when compared to no systemic therapy), suggesting that the association between the signature and overall survival may be similar in these groups. However, there was no significant association between survival and the signature score when conditioning on receiving non-platinum based systemic therapy or when not receiving systemic treatment (Table 2, Figure 5). There was also no significant association between survival and systemic treatment when the signature score was set to zero (Table 2). The models R^2^ = 0.32 and C-index was 0.72 (SE = 0.02). The corresponding clinical base model had a C-index of 0.71 (SE = 0.02) and R^2^ = 0.30.

**Figure 5.**
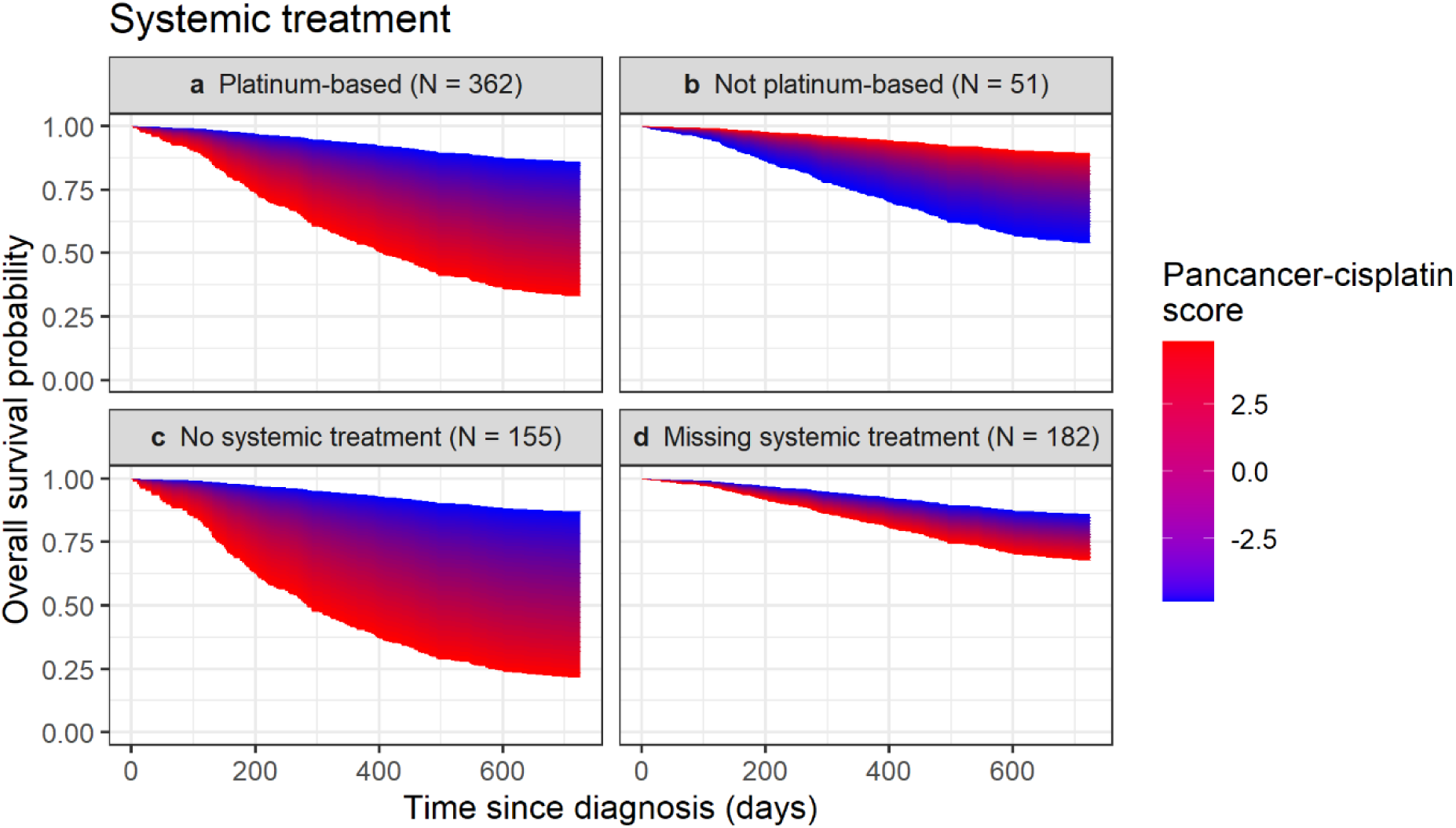
Estimated relationship between the *pancancer-cisplatin* signature and 2-year overall survival. **a** Relationship in patients that received platinum-based chemotherapy. **b** Relationship in patients that received non-platinum based systemic treatment. **c** Relationship in patients that did not receive systemic treatment. **d** Relationship in patients with missing information about systemic treatment. The signature score is scaled to a mean of 0 and a standard deviation of 1. Note the difference in sample size (N) per group.

In contrast to OS, we found no significant association between 2-year DFS and the *pancancer-cisplatin* signature (Table 2). There was also no evidence of interactions between the score and systemic treatment when platinum-based chemotherapy was the reference (p = 0.258 when compared to non-platinum based, p = 0.571 when compared to no systemic treatment). The models C-index was 0.70 (SE = 0.02) and R^2^ = 0.28. The C-index of the corresponding clinical base model was 0.70 (SE = 0.02) and R^2^ = 0.26. See Table S8 for details about other covariates included in the *pancancer-cisplatin* models.

#### 3.2.5. Predictive and prognostic signature: *Cl3-hypoxia*

The *Cl3-hypoxia* signature was significantly associated with 2-year OS in patients receiving non-cetuximab-based systemic treatments, where increasing the signature score resulted in a lower hazard (HR = 0.69 [CI: 0.54, 0.87], Table 2, Figure 6). A similar, but non-significant, association between OS and the GS was found for cetuximab-treated patients (HR = 0.67 [CI: 0.39, 1.14], Table 2, Figure 6). In contrast, an opposite association was found between OS and the signature in patients who did not receive systemic treatments, where higher signature scores were associated with higher hazards (HR = 1.70 [CI: 1.12, 2.58], Table 2, Figure 6).

**Figure 6.**
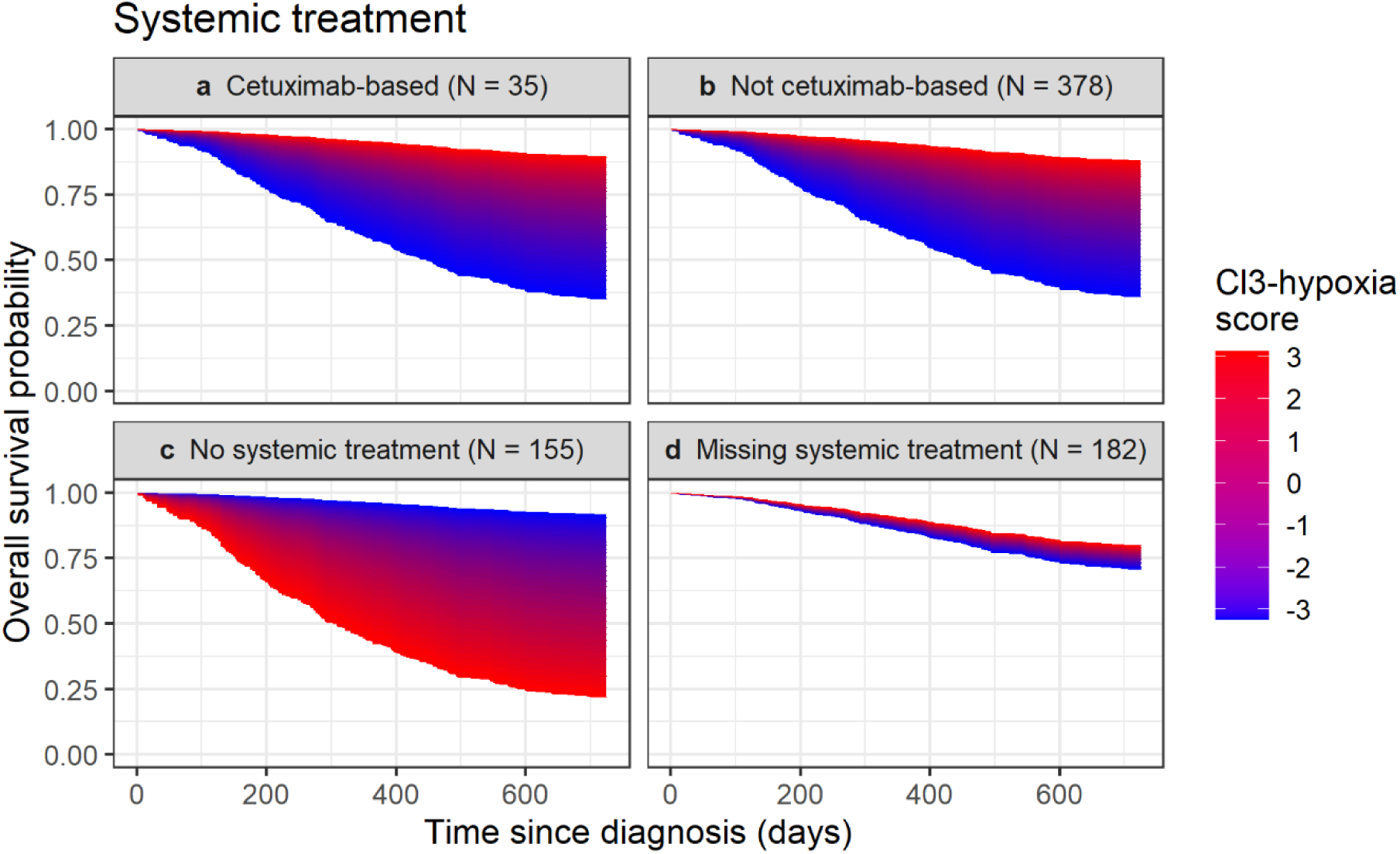
Estimated relationship between the *Cl3*-*hypoxia* signature and 2-year overall survival. **a** Relationship in patients that received cetuximab-based treatment. **b** Relationship in patients that received non-cetuximab based systemic treatment. **c** Relationship in patients that did not receive systemic treatment. **d** Relationship in patients with missing information about systemic treatment. The signature score is scaled to a mean of 0 and a standard deviation of 1. Note the difference in sample size (N) per group.

There was evidence of interactions between the signature and systemic treatment when using no systemic treatment as the baseline (comparison with cetuximab group: p = 0.006; comparison with non-cetuximab: p < 0.001), but no evidence of an interaction when comparing cetuximab-treated patients with patients receiving other types of systemic treatment (p = 0.919). When the GS score was set to zero, there was no significant association between survival and systemic treatment (Table 2). The models C-index was 0.73 (SE = 0.02) and R^2^ = 0.34. The C-index of the corresponding clinical base model (i.e. not including the GS as a covariate) was 0.71 (SE = 0.02) and R^2^ = 0.29.

Similar effect sizes and significance levels were found when analyzing 2-year DFS (Table 2). Increasing the signature score was associated with a reduced hazard in patients receiving systemic treatment, while being significantly associated with an increased hazard in patients not receiving systemic treatments (Table 2). There was evidence of interactions between the signature and systemic treatment when no systemic treatment was the baseline (comparison with cetuximab group: p = 0.006; comparison with non-cetuximab: p < 0.001), but no evidence of an interaction when comparing patients receiving cetuximab with patients receiving non-cetuximab therapies (p = 0.943). The models R^2^ = 0.32 and C-index = 0.71 (SE = 0.02). The C-index of the corresponding clinical base model was 0.70 (SE = 0.02) and R^2^ = 0.27. See Table S9 for details about other covariates included in the *Cl3-hypoxia GS*.

### 3.3. Sensitivity analyses

#### 3.3.1. Coding of treatment

We performed sensitivity analyses to evaluate the effect of using the received treatment as a proxy of intended treatment (Appendix 1). Results of these analyses are summarized below:

i. All treated patients started treatment within 6 months after diagnosis, with most starting treatment within 3-4 months. In the first 6 months after diagnosis, the mortality was approximately 3%.
ii. We obtained overlapping and similar treatment coefficients when comparing intended and received treatment in an external dataset with similar patients’ characteristics.
iii. We obtained overlapping and similar treatment coefficients when comparing received treatment with time-dependent treatment in a subset of patients where the timing of surgery was known.
iv. A landmark analysis showed overlapping and similar treatment coefficients as when using received treatment.
v. Quantitative bias analyses showed bias-adjusted estimates of treatment effects that overlap with estimates obtained when using received treatment.

#### 3.3.2. 5-year endpoints

Effect sizes and significance levels from models with 5-year OS and 5-year DFS were very similar to the 2-year survival models (see Figures S4-S13 for forest plots and Tables S5-S9 for detailed results). The main difference was in the model of 5-year OS and the *Cl3-hypoxia* signature, where there was no longer evidence of an association between the score and survival in patients not receiving systemic treatments (HR = 1.43 [CI: 0.96, 2.13], p = 0.076). Moreover, there was no longer evidence of an interaction between the score and systemic treatment when comparing patients untreated with systemic agents with patients receiving cetuximab (p = 0.068). The median follow-up time was 1325 days (approx. 3.63 years).

## 4. Discussion

In this study, we externally validated five gene signatures using one of the world’s largest collections of HNSCC patients with available gene expression, harmonizing and combining high quality clinical data from different studies. The development of this wide European dataset has shown that the integration of multisource clinical and biological databases is feasible. This successful integration is unprecedented and amplifies the knowledge about the complexity and the heterogeneity of HNSCCs, contributing to their qualification and quantification. The external validation ensures the robustness of the GSs and their potential generalizability. In addition, tumor heterogeneity and complexity coupled with the variety of treatment options imposes a biology-driven personalized approach in the curative setting of HNSCC patients. Unlike more frequent tumors (e.g., breast cancer, non-small cell lung cancer, or colorectal cancer), GSs have not been included yet either in clinical decision making, or in clinical trial eligibility and stratification of HNSCC patients.

Except for the model testing associations between *RSI* and disease-free survival, all models including gene signatures outperformed their corresponding clinical omics-free base models, as indicated by C-index and/or R^2^. Our results confirmed two potentially prognostic signatures (*172-GS* and *3 clusters HPV*) and validated one potentially prognostic/predictive signature (*Cl3-hypoxia*) for cetuximab- and chemosensitivity. However, the results for two potentially predictive signatures (*RSI* and *pancancer-cisplatin*) were less conclusive, despite showing associations with overall survival in patients receiving radiotherapy or platinum-based chemotherapy.

The signature *172-GS*^18^ was validated as a prognostic indicator for both overall and disease-free survival in patients with HPV-unrelated disease. A higher signature score is associated with a higher risk. The signature may also be prognostic in HPV-positive oropharyngeal cancer patients, as given by the lack of evidence of an interaction between the signature and HPV-status. This contrasts with previously published results^19^, possibly due to adjustments for covariates and uncertainties in estimates of the signature effect for HPV-positive patients.

The *3 clusters HPV* signature^20^ was validated as prognostic for both OS and DFS in HPV-positive oropharyngeal cancer patients, consistent with a previous validation study^19^.

Additionally, there was no evidence of the signature being prognostic for HPV-negative patients, suggesting its relevance specifically to HPV-positive oropharyngeal cancer.

In our study, we found a link between the radiosensitivity index (*RSI*)^21^ and overall survival in patients who underwent radiotherapy, consistent with other studies^18,19,22,65^. However, we did not observe any interaction between *RSI* and radiotherapy. We also found a similar but not significant association between *RSI* and OS in patients who did not receive radiotherapy. This suggests that *RSI* may be more prognostic for survival than predictive of radiosensitivity. This surprising finding of a prognostic effect of *RSI* in non-irradiated patients may be because of the relatively low number of non-irradiated patients in our study. In contrast to other studies^22,66^, we did not find a connection between *RSI* and DFS.

To our knowledge this is the first external validation of the *pancancer-cisplatin* signature^23^ in head and neck cancer patients. We found a negative association between the signature score and overall survival (OS) in patients receiving platinum-based chemotherapy. Given that higher GS scores were linked to higher cisplatin-sensitivity in the original publication^23^, our finding is in the opposite direction of what is expected. Additionally, we did not find evidence of an interaction between systemic treatment and the signature. Due to an imbalance in the number of patients with each type of systemic treatment, we could not draw definitive conclusions about the potential prognostic or predictive effects of the signature. Lastly, we found no associations between disease-free survival (DFS) and the *pancancer-cisplatin* signature, suggesting that its predictive power may vary with clinical outcomes and not be universal for all cancers.

In general, a GS is prognostic if it is able to forecast patient survival independently of the received treatment. At the same time, if a prognostic GS is tested on a subset of patients receiving curative therapies including a specific approach, then the prognostic performance necessarily implies a certain predictive capability. In this scenario, the *Cl3-hypoxia*^24^ signature score was positively associated with both OS and DFS in patients receiving systemic treatment but negatively associated with survival in subjects not receiving systemic agents, suggesting the signature is predictive of sensitivity to systemic therapy. A similar positive association with survival endpoints was found in both cetuximab-treated patients and in patients with chemotherapy, which did not include anti-EGFR agents. The association between signature and survival is consistent with previous findings in HPV-positive oropharyngeal cancer^19^ and in oral premalignant lesions^25^. Previous studies have suggested that the signature is related to cetuximab-sensitivity^26,27^, but our results indicate that it is predictive of sensitivity to systemic treatment in general.

This study offers various opportunities for further improvement and refinement. Firstly, we used the received treatment as a proxy for intended treatment. While this breaks an assumption of the Cox regression model, we performed extensive sensitivity analyses that suggest the models are robust to this violation. Secondly, 3-7% of patients (depending on the model) were excluded for missing overall survival status or timing, and 6-19% were excluded for missing DFS. This complete case exclusion reduced the sample sizes. Still, because the missingness was primarily structural^58^, where the survival endpoint was missing for all patients in a study, it is unlikely that it biased the results. Third, we used the missing indicator method for missing covariates (with missingness ranging from 3-30% depending on the covariate), which can lead to biased results in non-randomized studies^67^, but is unlikely to result in bias unless the covariate is a strong confounder or missing in extreme proportion (>50% missing)^68^. Moreover, like the missingness in endpoints, the missingness in covariates was primarily structural, and alternative regression-based imputation methods were therefore not a viable option. Lastly, the imbalance in the number of patients with different treatments (e.g., only 35 patients received cetuximab) made it difficult to precisely estimate interactions and effects of gene signatures in small groups and made conclusions less clear.

We externally validated five gene signatures using a large integrated dataset of HNSCC patients. Our results validated two prognostic signatures, *172-GS* and *3 clusters HPV* signature, in HPV-negative patients and HPV-positive patients, respectively. We also validated that the potentially predictive signatures *RSI* and *pancancer-cisplatin* were prognostic of survival in patients with radiotherapy and platinum-based chemotherapy, respectively, but could not conclude if these signatures are also predictive of radiosensitivity or platinum-chemosensitivity. Lastly, we validated the *Cl3-hypoxia* signature as predictive of sensitivity to systemic treatment, making this signature a good candidate for use in personalized treatment decisions.

Many clinical studies aimed at intensifying^69–71^ or de-escalating^72–74^ standard treatments have failed in head and neck oncology. A possible drawback could be the lack of an appropriate patient selection, which may have hampered the opportunity to detect a signal of clinical benefit with a given therapeutic approach. The significant biological differences in the current study demonstrate the high heterogeneity of HNSCCs. Therefore, future clinical trials should pursue a better patient selection that includes a broader use of GS and incorporates them as inclusion criteria or stratification factors. In this scenario, the results of our external validation in one of the widest HNSCC datasets may be considered a historical benchmark for the design of future studies.

## 5. Data availability

The SuPerTreat dataset is hosted by the Services for Sensitive Data (TSD) at the University of Oslo. Anonymized data containing only the survival endpoints and gene signature scores will be uploaded to Zenodo prior to publication. Access to the original data may be granted by the data owners upon application.

## 6. Code availability

The underlying code (R scripts) used to analyse and validate gene signatures is available on GitHub and can be accessed via this link https://github.com/erlendfossen/SuPerTreat_GS_validation.

## 7. Acknowledgements

This work is part of the research project “Supporting Personalized Treatment Decisions in Head and Neck Cancer through Big Data (SuPerTreat)” funded within ERA PerMed JTC2019 Joint transnational call for proposals (2019) for “Personalised medicine: multidisciplinary research Toward implementation”, research grant nr. ERAPERMED2019-281 and supported by FRRB (Fondazione Regionale per la Ricerca Biomedica), BMBF (Bundesministerium für Bildung und Forschung), RCN (Norges forskningsråd), ANR (Agence nationale de la recherche) and GSRT (General Secretariat for Research and Technology). MM-S received funding from the European Union’s Horizon 2020 Research and Innovation program under the Marie Skłodowska-Curie Actions Grant, agreement No. 80113 (Scientia fellowship). BD2Decide received funding from the European Union Horizon 2020 Framework Programme, Grant/Award Number: 689715. We thank the researchers, clinicians and other staff that designed and conducted the studies that this work builds on. We also thank the head and neck cancer patients that took part in the studies.

## 8. Author contributions

Conceptualization: E.I.F.F., M.M.S., M.L., A.F., I.T., C.L.T., L.L, L.D.C, S.C.; Data curation: E.I.F.F., M.M.S., L.L.P, E.E.P, L.H, L.D.C, S.C.; Formal analysis: E.I.F.F., M.M.S., M.L., A.F., L.D.C, S.C.; Funding acquisition: A.F., E.H., M.F.C.U, G.F., I.T., L.L.; Investigation: I.T., V.S., K.S., C.L.T., M.K., S.T., M.P., L.L, L.D.C, S.C.; Methodology: E.I.F.F., M.M.S., M.L., A.F., L.L, L.D.C, S.C.; Resources: A.F., L.L.; Software: E.I.F.F., M.M.S.; Supervision: M.L., A.F., M.F.C.U., G.F.; Visualization: E.I.F.F.; Writing-original draft: E.I.F.F.; Writing-review & editing: All authors. All authors approved the submission of this manuscript. L.D.C. and S.C. contributed equally to this work as co-last authors.

