## Supplementary Appendices and Figures for "External validation of prognostic and predictive gene signatures in head and neck cancer patients"

#### Appendix 1: Sensitivity analyses of treatment

Sensitivity analyses were performed to investigate how sensitive results are to using received treatment (surgery, systemic treatment and radiotherapy) as a proxy for intended treatment in Cox regression models.

**Mortality before start of treatment**

We had information about the timing of treatments in a small subgroup of patients (N = 80). In these data, 90% of patients underwent surgery within 3 months after diagnosis and all had undergone surgery within 6 months (180 days). Approximately 85% of patients that received radiotherapy or systemic treatment started therapy within 4 months (120 days), with all starting within 180 days. Moreover, during data harmonization, we made sure that any treatment starting many months after diagnosis was considered treatment of cancer recurrence and thus not part of our curative intent treatment covariates. Lastly, clinicians involved in data collection confirmed that all patients followed a standard of care where they started treatment within 180 days, and often much earlier than this.

Mortality during the first 180 days after diagnosis was approximately 5% (Fig. A1A). However, this is an upper limit because most patients start treatment after only 30-90 days and because the survival curve included some patients that died after already having started their intended treatment. To account for patients that died after having received treatment, we conducted a competing risk analysis on a subset of patients (N = 437) where the timing of surgery was known. In the competing risk model, death and surgery were treated as competing events and we estimated the cumulative incidence of each event in the first 180 days after diagnosis. From this analysis we see that mortality was approximately 3% and that most patients that underwent surgery did so within 60 days in the BD2_UDUS study and within 120 days in the BD2_INT_MI study (Fig. A2).

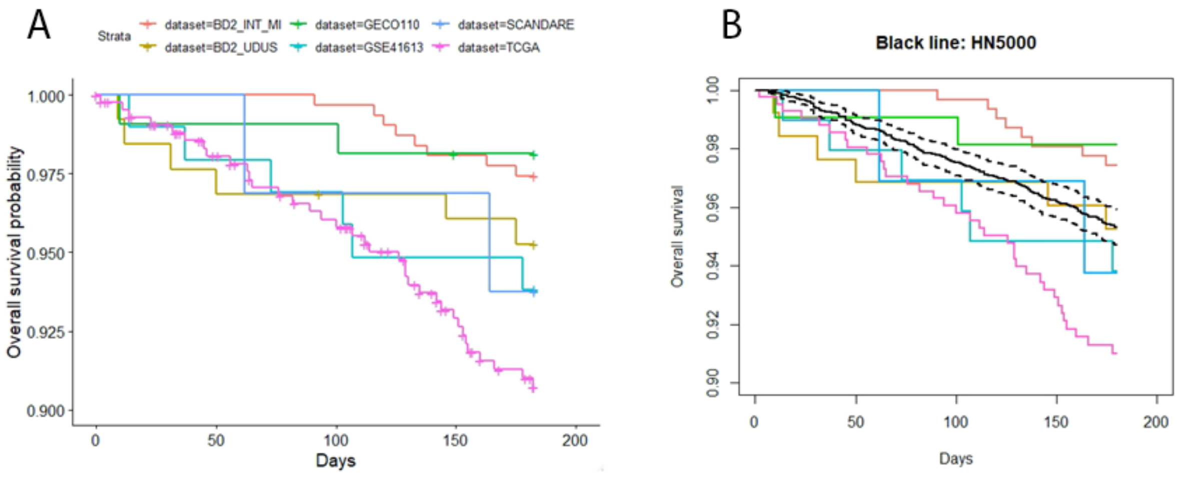

**Figure A1**. Overall survival from diagnosis to 180 days. Colors represent different datasets/studies. Pluss signs (+) indicate censoring. **A)** Kaplan-Meier curves for 6 datasets included in clinical scenarios where gene signatures were tested. **B)** Same as in A, but the black line represents the survival curve of an external dataset, H&N5000, with dashed lines showing 95% confidence interval for the H&N5000 study.

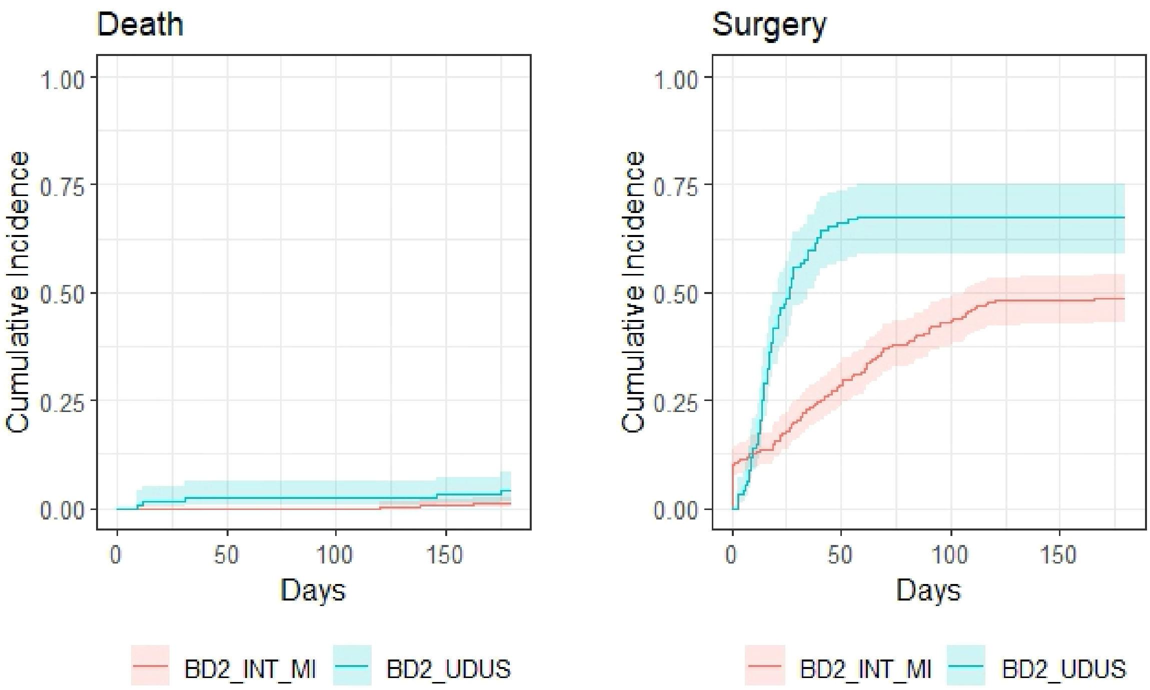

**Figure A2**. Cumulative incidence of deaths (left panel) and surgery (right panel) for two datasets (BD2_INT_MI, BD2_UDUS) in SuPerTreat. Shaded area represents 95% confidence intervals. Cumulative incidence was obtained from a competing risk model with death and surgery as competing risks.

**Intended versus received treatment**

We used data from an external study, Head and Neck 5000 (H&N5000) (Ness *et al.* (2014); *Ness et al.* (2016)), to compare coefficients from two Cox regression models, one model with received treatment and one model with intended treatment. H&N5000 is a large prospective clinical cohort study that has recruited 5511 people with head and neck cancer from 76 centres across the UK. The study is sponsored by University Hospitals Bristol and Weston NHS Foundation Trust and is one of the largest studies of its type for head and neck cancer patients. The study has captured several hundred variables, including clinical and demographic characteristics, but did not contain any gene expression data.

We applied the same eligibility criteria, except the requirement of having gene expression data, that was used in SuPerTreat to the H&N5000 data and ended up with N = 4486 eligible patients. H&N5000 patients had overlapping survival curves with patients from SuPerTreat (Fig. A1B). A total of 21% of H&N5000 patients received a different treatment than intended/planned at baseline diagnosis, with the biggest contribution from patients that had no intended radiotherapy but ended up receiving radiotherapy (36% of patients with no intended radiotherapy received radiotherapy). For SuPerTreat data we could get a rough estimate of how many patients deviated from treatment plans based on two clinical rules of thumbs: i) TNM stages I and II should have unimodal treatment. Cases with multimodal treatment are considered deviations in treatment; ii) TMN stages III, IVA and IVB should have multimodal treatment. Cases with unimodal treatment are considered deviations in treatment. Based on these rules of thumb, we estimated that 15% of patients deviated from intended treatment. This is likely an underestimate since the rules of thumbs cannot identify cases that kept the same treatment modality but changed which specific treatment was received (e.g. receiving chemotherapy instead of intended radiotherapy).

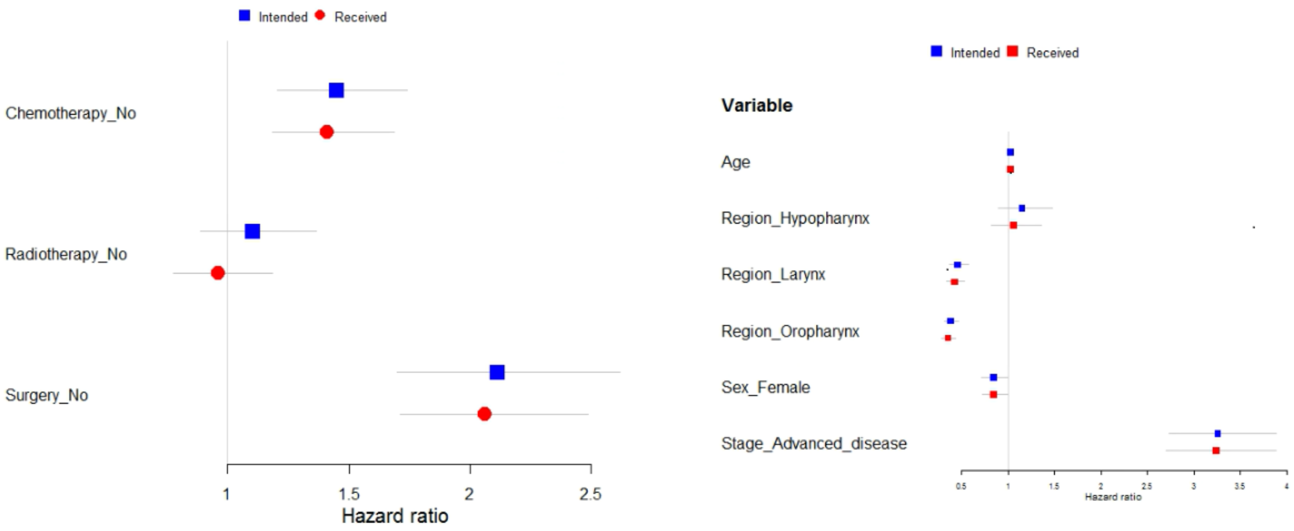

**Figure A3**. Hazard ratios (and 95% confidence intervals) from two Cox regression models with overall survival censored at 2 years as the endpoint, one including intended treatment (blue squares) and one including received treatment (red circles). Left panel shows treatment coefficients where the reference was "Yes". Right panel shows coefficients of other covariates, with references being oral cavity for tumor region, male for sex, and early disease for tumor stage.

Using H&N5000 data, two Cox proportional hazard models were built with overall survival censored at 2 years as the endpoint, and age, sex, tumor region, cTN disease extension and treatments (chemotherapy, radiotherapy, surgery) as covariates. The difference between the models was that one model included the intended treatments while the other included the received treatments. When comparing estimated hazard ratios from the two models we find overlapping confidence intervals (Fig. A3), suggesting that received treatment was a good proxy for intended treatment.

**Time-dependent treatment**

We knew the timing of surgery for a subset of SuPerTreat patients (N = 437). For this subset of patients, we built two models with overall survival censored at 2 years as the endpoint, one Cox regression model with received treatment (codes as a baseline characteristic) and one modified Cox regression model with surgery coded as time-dependent (switching from "No" to "Yes" at an individual's time of surgery). Other covariates were gene signature *172-GS*, chemotherapy (received or not), age, sex, TNM stage, dataset, radiotherapy (received or not), smoking and tumor region. When comparing estimated hazard ratios from the two models we find overlapping confidence intervals (Fig. A4). Similar results were found when using disease-free survival censored at 2 years as the endpoint (not shown).

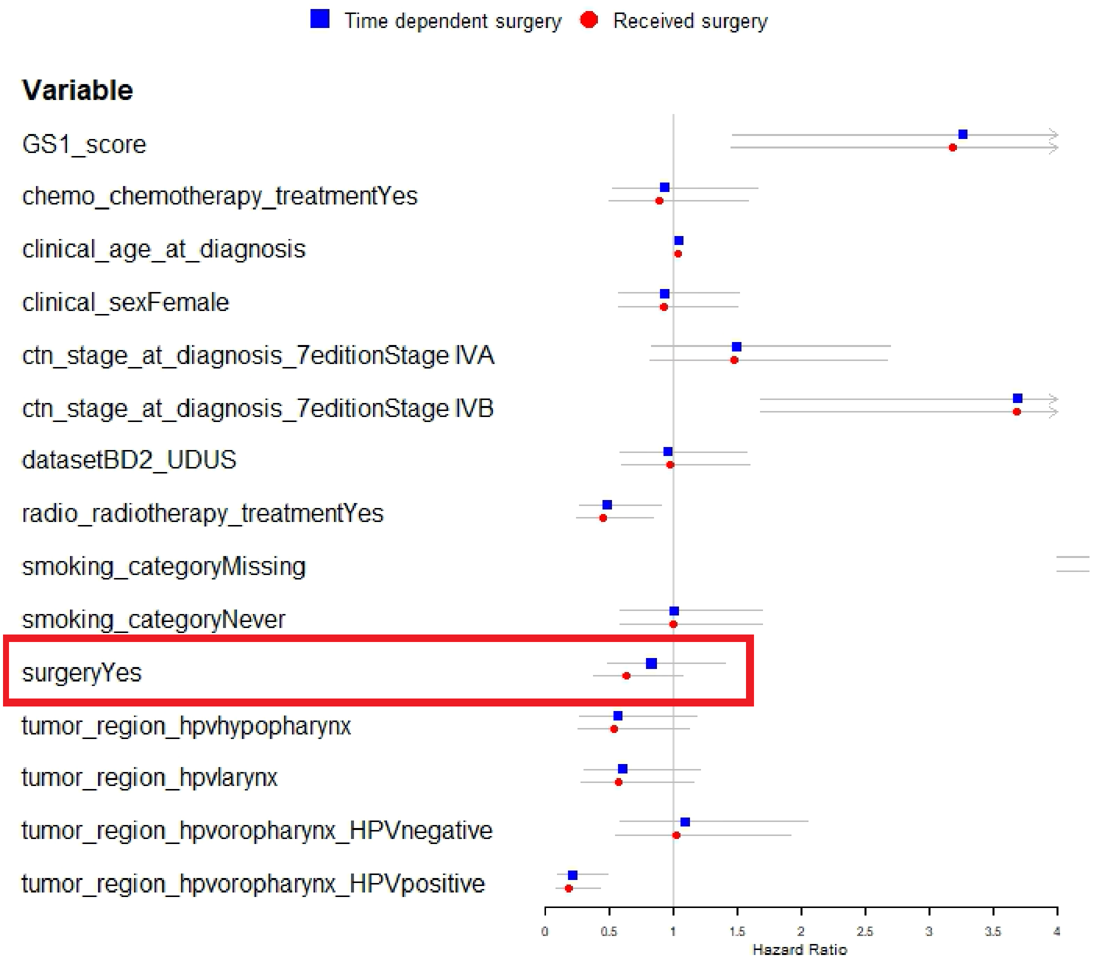

**Figure A4**. Hazard ratios (and 95% confidence intervals) from two Cox regression models with overall survival censored at 2 years as the endpoint, one modeling surgery as time-dependent (blue squares) and one including received treatment (red circles). The surgery coefficient is highlighted in red. GS1_score represents the signature score for gene signature *172-GS*. References for categorical variables: no chemotherapy, male, Stage III, BD2_INT_MI dataset, no radiotherapy, current or former smoking, no surgery, oral cavity for tumor region. Arrow on error bars imply that the upper confidence boundary is outside the range of values showed on the plot. Point estimates for smoking missing category were HR = 15 for received surgery model and HR = 16 for time-dependent model, with large overlapping confidence intervals.

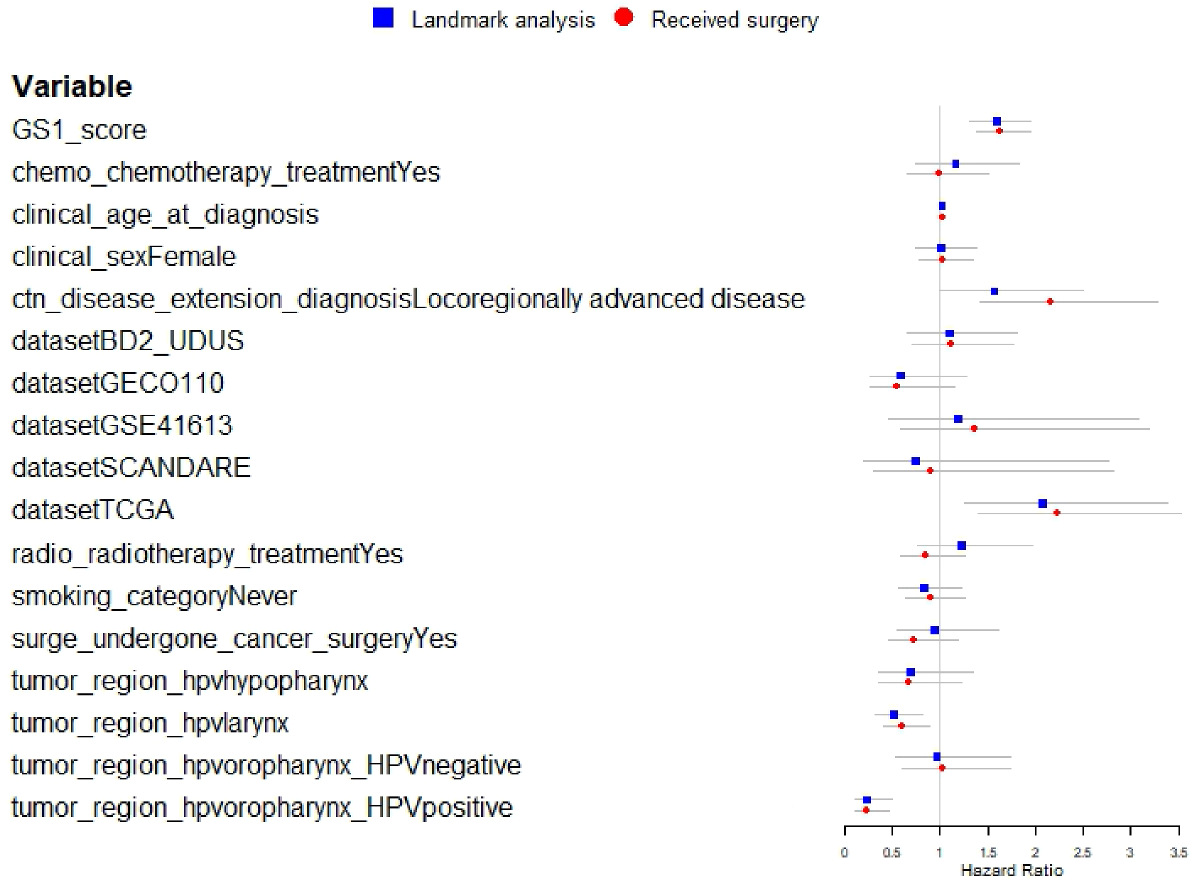

**Figure A5**. Hazard ratios (and 95% confidence intervals) from two Cox regression models with overall survival censored at 2 years after diagnosis as the endpoint, one model following patients from 180 days after diagnosis (blue squares) and one model following patients from the day of diagnosis (red circles). GS1_score represents the signature score for gene signature *172-GS*. References for categorical variables: no chemotherapy, male, early disease, BD2_INT_MI dataset, no radiotherapy, current or former smoking, no surgery, oral cavity for tumor region.

**Landmark analysis**

A landmark analysis involves choosing a fixed time point after cohort entry, a landmark, and excluding any patients that had an event or were censored prior to this time point. All remaining patients are then followed from the landmark and hazard ratios are estimated. By choosing a landmark time point after all patients should have started treatment, we can remove or greatly reduce immortal time bias. We fit two Cox models and compared hazard ratio estimates of these: i) a model that corresponds to the model used to test the *172-GS* signature (N = 1097), where 2-year overall survival was the endpoint, and the signature score, received treatments (chemotherapy, radiotherapy and surgery), age, sex, cTN disease extension, dataset/study ID, tumor region and HPV status, and smoking status were covariates; ii) Landmark model (N = 976), where patients that had an event or were censored the first 180 days after diagnosis were excluding and the remaining patients were followed from day 181 until 2 years after diagnosis. 180 days was used as the landmark since all patients would start treatment within this time. The landmark model included the same covariates as the previous model. When comparing estimated hazard ratios from the two models we find overlapping confidence intervals (Fig. A5).

**Quantitative bias analyses**
We performed summary-level quantitative bias analyses (QBA) to adjust for bias resulting from misclassification of intended treatments. When the received treatment does not match the intended treatment, this can be considered misclassification of the treatment. In summary-level QBA we use estimates of bias parameters (sensitivity and specificity) to adjust our observed data to represent bias-adjusted data and use these adjusted data to estimate bias-adjusted effect size of treatments.

We obtained estimates of sensitivity (probability of being correctly classified as “Yes” for intended treatment) and specificity (probability of being correctly classified as “No” for intended treatment) from the external H&N5000 dataset (described earlier) where both intended and received treatment was known. We expected to see differential misclassification where patients in worse condition were more likely to deviate from planned treatment and thus more likely to be misclassified than patients with a good prognosis. To account for this, we stratified the data based on survival status 2 years after diagnosis and estimated sensitivity and specificity separate for these two groups. We observed high sensitivity (≥ 0.9) for all treatments (Table A1), meaning that patients that had planned a specific treatment also tended to receive that treatment. Specificity was lower (Table A1), especially for radiotherapy, meaning that patients without planned radiotherapy often ended up receiving radiotherapy.

We stratified our SuPerTreat data by survival/event status at 2 years after diagnosis and obtained the number of patients with a given treatment and survival/event status. Using these observed data (Table A2) we estimated an odds ratio for each treatment by using univariate logistic regression. We also obtained a bias-adjusted odds ratio for each treatment by using the bias parameters from Table A1 to adjust the observed data and then using univariate logistic regression. This was done for both overall survival and for disease-free survival. The results (Table A3) show that all bias-adjusted odds ratios fall within the confidence intervals of the unadjusted odds ratios, suggesting that the bias is small.

**Table A1**. Sensitivity and specificity estimates for intended treatments. Bias parameters were estimated in H&N5000 data where both the intended and received treatment was known.

| **Treatment** | **Survival status 2 years after diagnosis** | **Sensitivity** | **Specificity** |
|---|---|---|---|
| Chemotherapy | Alive | 0.95 | 0.90 |
| | Dead | 0.90 | 0.86 |
| Radiotherapy | Alive | 0.98 | 0.68 |
| | Dead | 0.93 | 0.48 |
| Surgery | Alive | 0.95 | 0.97 |
| | Dead | 0.92 | 0.98 |

**Table A2**. Observed number of patients for different combinations of treatments and survival/event status 2 years after diagnosis. OS = overall survival. DFS = disease-free survival. CT = chemotherapy. RT = radiotherapy. Surg = surgery.

| Endpoint status | CT = yes | CT = no | RT = yes | RT = no | Surg = yes | Surg = no |
|---|---|---|---|---|---|---|
| OS alive | 442 | 369 | 733 | 215 | 786 | 215 |
| OS dead | 170 | 129 | 287 | 79 | 299 | 90 |
| DFS no event | 379 | 262 | 585 | 148 | 606 | 175 |
| DFS event | 197 | 162 | 314 | 71 | 291 | 130 |

**Table A3**. Unadjusted and bias-adjusted treatment effects on two survival endpoints estimated with univariate logistic regression. The reference was "Yes" for all intended treatments.

| Endpoint | Treatment | Odds ratio [95% CI] | Bias-adjusted odds ratio [95% CI] |
|---|---|---|---|
| Overall survival | Chemotherapy | 0.91 [0.70, 1.19] | 0.85 [0.65, 1.11] |
| | Radiotherapy | 0.83 [0.61, 1.13] | 0.97 [0.75, 1.27] |
| | Surgery | 1.10 [0.83, 1.46] | 0.93 [0.68, 1.26] |
| Disease-free survival | Chemotherapy | 1.19 [0.92, 1.54] | 1.18 [0.91, 1.53] |
| | Radiotherapy | 0.89 [0.65, 1.22] | 1.02 [0.77, 1.34] |
| | Surgery | 1.55 [1.19, 2.02] | 1.46 [1.10, 1.94] |

#### Appendix 2: Gene signature details

***172-GS* signature**
Mean and SD of the signature score on original scale:

- Overall survival data: mean = 1.08, SD = 0.701
- Disease-free survival data: mean = 0.513, SD = 0.735

Genes (given as gene symbols) that are part of the signature, but were missing in our data:

- Overall survival data: SH3RF3, CENPBD1, ZC3H12B, GVINP1, PCGF2, ST7-AS1, TMC01-AS1, POFUT1, TMC8, ARMCX4, SLFN5, RASSF5, NPIPB5, XKR6, SNX29P2, CPNE9, CHSY3, FAM 53C, FGD2, KCNE 5, RSPH6A, LOC153684, UBAC2, PHETA2, LILRA6, PLEKHM3, METTL7B, FSTL1, SCFD2, c11orf49, FXYD2, NAP1L5, DOC2B, PALM2AKAP2, KIR2DS2, KEL, ARHGAP30, STBD1, DAPK3, GPRASP2, KLRC2, LSP1P5
- Disease-free survival data: SH3RF3, CENPBD1, ZC3H12B, GVINP1, ST7-AS1, TMC01-AS1, POFUT1, TMC8, ARMCX4, SLFN5, NPIPB5, XKR6, SNX29P2, CPNE9, CHSY3, FAM 53C, FGD2, KCNE 5, LOC153684, UBAC2, PHETA2, PLEKHM3, METTL7B, FSTL1, SCFD2, c11orf49, NAP1L5, PALM2AKAP2, KIR2DS2, ARHGAP30, STBD1, DAPK3, GPRASP2, LSP1P5

***3 clusters HPV* signature**
Mean and SD of the signature score on original scale:

- Overall survival data: mean = -36.658, SD = 3.473
- Disease-free survival data: mean = -38.752, SD = 3.407

Genes (given as gene symbols) that are part of the signature, but were missing in our data:

- Overall survival data: FCER2, IKBKG, IGHVII-44-2, BPIFA3, ZNF300P1, DNAJB8, FLCN, LINC00927, EPHA1-AS1, LINC00226, FKBP9P1, BEND4, MIR30E, TRIM51CP, GAPLINC
- Disease-free survival data: IGHVII-44-2, ZNF300P1, DNAJB8, FLCN, LINC00927, EPHA1-AS1, LINC00226, FKBP9P1, BEND4, MIR30E, TRIM51CP, GAPLINC

**Radiosensitivity index (*RSI*) signature**
Mean and SD of the signature score on original scale:

- Overall survival data: mean = 0.116, SD = 0.195
- Disease-free survival data: mean = 0.110, SD = 0.196

***Pancancer-cisplatin* signature**
Mean and SD of the signature score on original scale:

- Overall survival data: mean = -0.612, SD = 0.519
- Disease-free survival data: mean = -0.612, SD = 0.489

Genes (given as gene symbols) that are part of the signature, but were missing in our data:

- Overall survival data: SLC25A45, TMC4
- Disease-free survival data: SLC25A45, TMC4

***Cl3-hypoxia* signature**
Mean and SD of the signature score on original scale:

- Overall survival data: mean = 9.404, SD = 3.877
- Disease-free survival data: mean = 9.408, SD = 3.777

Genes (given as gene symbols) that are part of the signature, but were missing in our data:

- Overall survival data: PLPP4, PEAR1, FIBIN
- Disease-free survival data: PLPP4, PEAR1, FIBIN

#### Appendix 3: Cox proportional hazard models

Formula for each of the Cox models described in the main text.

All follow the general Cox model equation:
$\lambda(t|X) = \lambda_0(t)\exp(\beta_1 X_1 + \beta_2 X_2 + \cdots + \beta_p X_p)$,
where t = time, $\lambda_0(t)$ is the baseline hazard, β is the estimated coefficient and X are realized values for p covariates.

OS = 2-year overall survival. DFS = 2-year disease-free survival.

***172-GS*** **models:**
OS: OS ~ age + sex + tumor region + smoking status + undergone surgery + received radiotherapy + received systemic treatment + study ID + cTN disease extension + HPV status × 172-GS score

DFS: DFS ~ age + sex + tumor region + smoking status + undergone surgery + received radiotherapy + received systemic treatment + study ID + cTN disease extension + HPV status × 172-GS score

***3 clusters HPV*** **models:**
OS: OS ~ age + sex + tumor region + smoking status + undergone surgery + received radiotherapy + received systemic treatment + study ID + cTN disease extension + HPV status × 3 clusters HPV score

DFS: DFS ~ age + sex + tumor region + smoking status + undergone surgery + received radiotherapy + received systemic treatment + study ID + cTN disease extension + HPV status × 3 clusters HPV score

***RSI*** **models:**
OS: OS ~ age + sex + tumor region and HPV status + smoking status + undergone surgery + received systemic treatment + study ID + cTN disease extension + received radiotherapy × RSI score

DFS: DFS ~ age + sex + tumor region and HPV status + smoking status + undergone surgery + received systemic treatment + study ID + cTN disease extension + received radiotherapy × RSI score

***Pancancer-cisplatin*** **models:**
OS: OS ~ age + sex + tumor region and HPV status + smoking status + undergone surgery + received radiotherapy + study ID + TNM stage + systemic treatment agent (platinum-based) × pancancer-cisplatin score

DFS: DFS ~ age + sex + tumor region and HPV status + smoking status + undergone surgery + received radiotherapy + study ID + TNM stage + systemic treatment agent (platinum-based) × pancancer-cisplatin score

***Cl3-hypoxia*** **models:**
OS: OS ~ age + sex + tumor region and HPV status + smoking status + undergone surgery + received radiotherapy + study ID + TNM stage + systemic treatment agent (cetuximab-based) × Cl3-hypoxia score

DFS: DFS ~ age + sex + tumor region and HPV status + smoking status + undergone surgery + received radiotherapy + study ID + TNM stage + systemic treatment agent (cetuximab-based) × Cl3-hypoxia score

### Supplementary figures

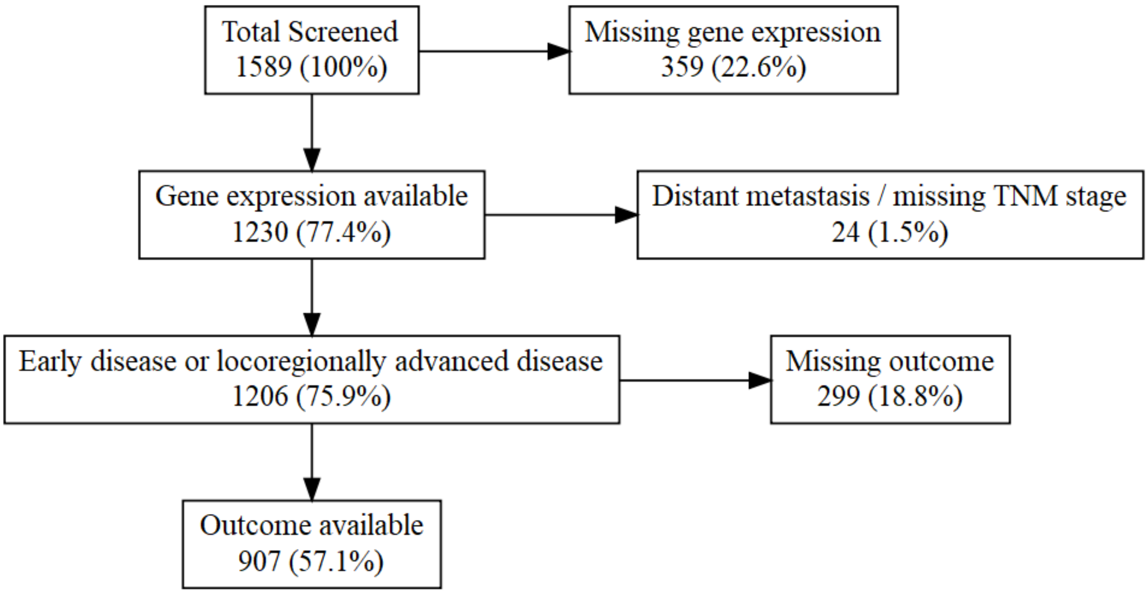

**Figure S1.** Exclusion flowchart for eligibility in models testing the signatures *172-GS*, *3 clusters HPV* and *RSI* with disease-free survival as the endpoint.

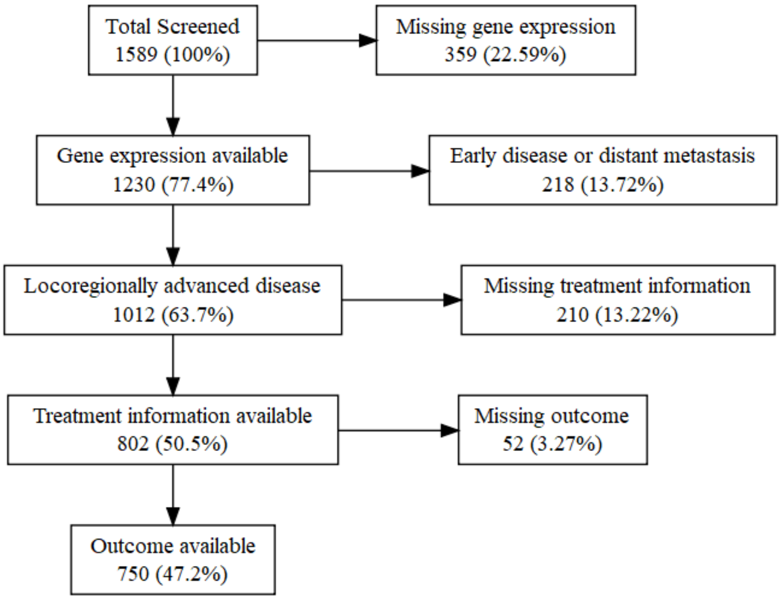

**Figure S2.** Exclusion flowchart for eligibility in models testing the signatures *pancancer-cisplatin* and *Cl3-hypoxia* with overall survival as the endpoint.

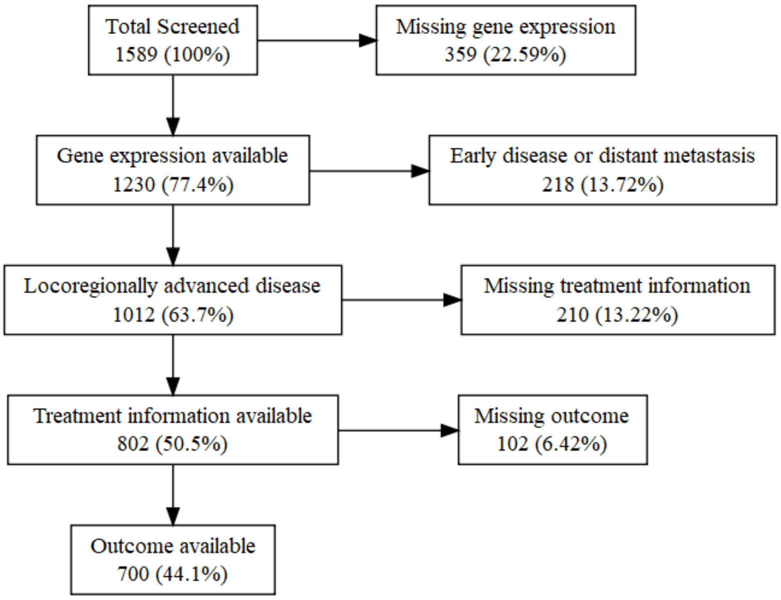

**Figure S3**. Exclusion flowchart for eligibility in models testing the signatures *pancancer-cisplatin* and *Cl3-hypoxia* with disease-free survival as the endpoint.

For **figures S4-S13**, the following reference levels were used for covariates that were in common: sex = male; dataset/study id = BD2_MI_INT; radiotherapy = no; smoking = never; surgery = no. Other reference levels are given in the figure captions.

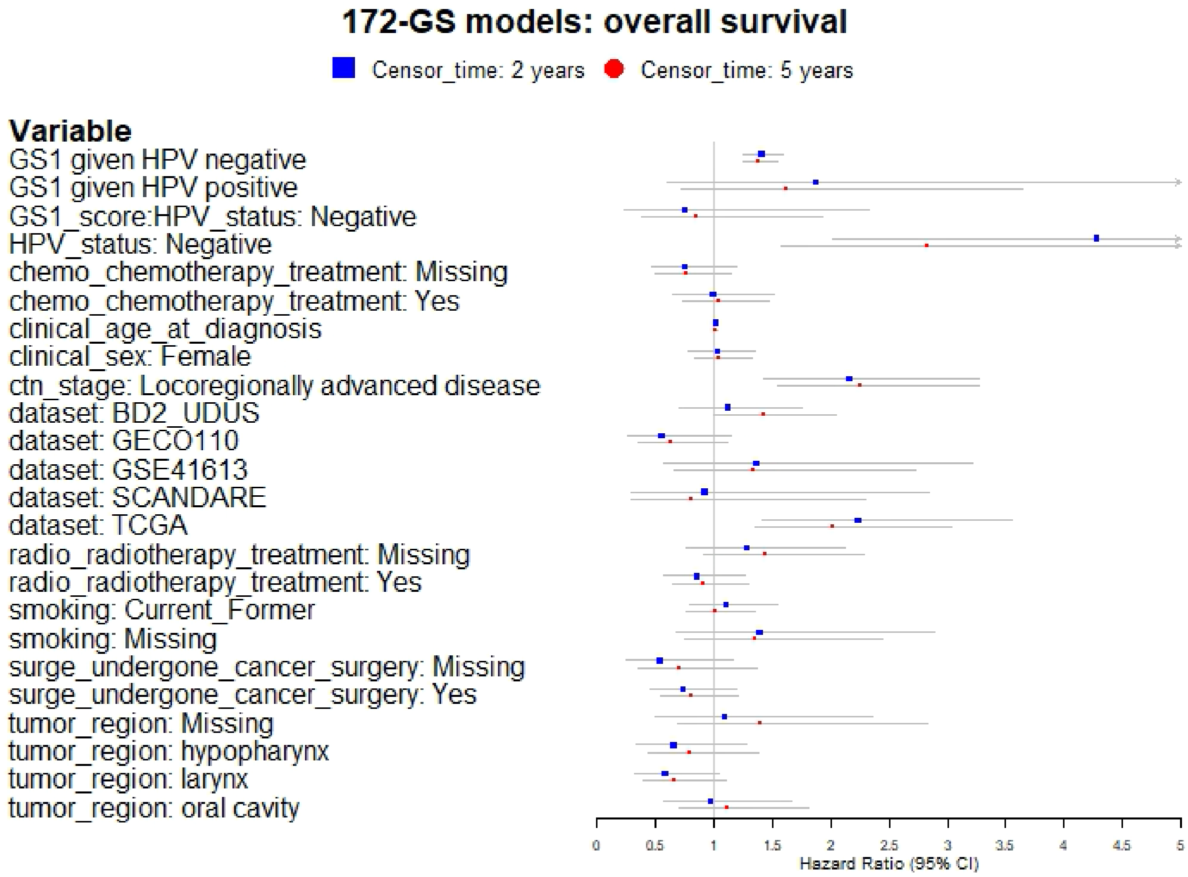

**Figure S4.** Forestplot showing the hazard ratio estimates (with error bars representing 95% confidence intervals) for different variables/parameters in multivariable models testing the *172-GS* signature where overall survival was the endpoint. Two models are compared, where the difference is the censoring time of the endpoint (2 years or 5 years since diagnosis). Arrow on error bars implies that the upper confidence boundary is outside the range of values showed on the plot (see Table S6 for precise estimates). Early disease is the reference for ctn_stage, oropharynx is the reference for tumor_region, HPV-positive is the reference for HPV_status, and no chemotherapy is the reference for chemo. GS1 is the gene signature score and GS1_score:HPV_status represents an estimated interaction between the signature score and HPV status.

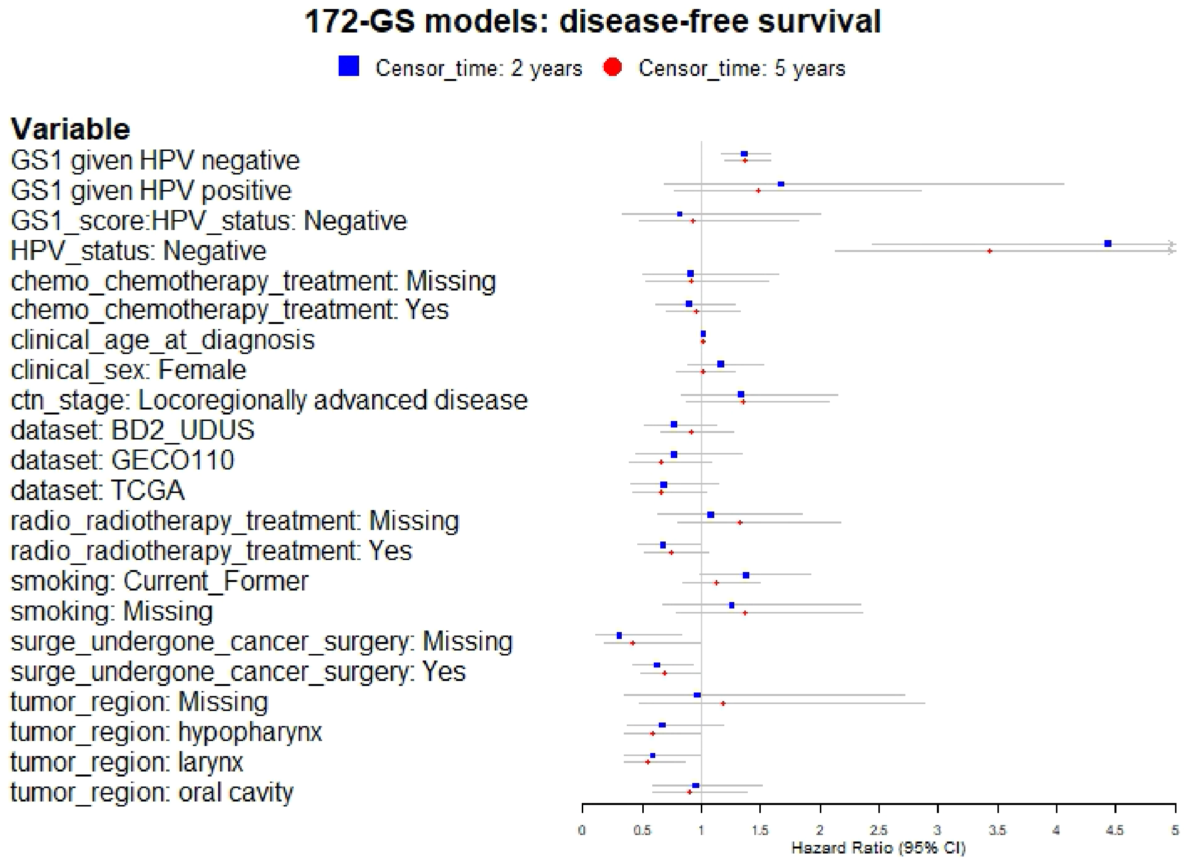

**Figure S5.** Forestplot showing the hazard ratio estimates (with error bars representing 95% confidence intervals) for different variables/parameters in multivariable models testing the *172-GS* signature where disease-free survival was the endpoint. Two models are compared, where the difference is the censoring time of the endpoint (2 years or 5 years since diagnosis). The arrow on error bars implies that the upper confidence boundary is outside the range of values shown on the plot (see Table S6 for precise estimates). Early disease is the reference for ctn_stage, oropharynx is the reference for tumor_region, HPV-positive is the reference for HPV_status, and no chemotherapy is the reference for chemo. GS1 is the gene signature score and GS1_score:HPV_status represents an estimated interaction between the signature score and HPV status.

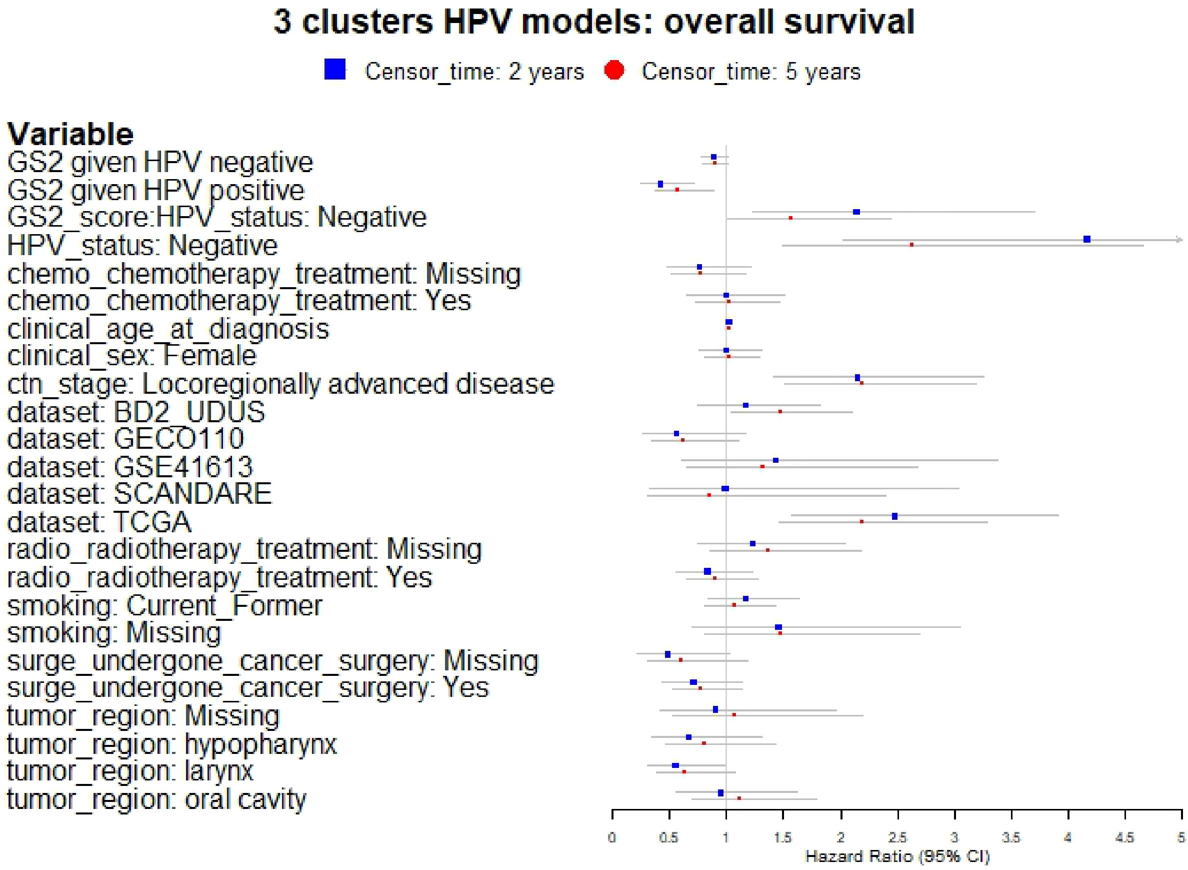

**Figure S6.** Forestplot showing the hazard ratio estimates (with error bars representing 95% confidence intervals) for different variables/parameters in multivariable models testing the *3 clusters HPV* signature where overall survival was the endpoint. Two models are compared, where the difference is the censoring time of the endpoint (2 years or 5 years since diagnosis). The arrow on the error bars implies that the upper confidence boundary is outside the range of values showed on the plot (see Table S7 for precise estimates). Early disease is the reference for ctn_stage, oropharynx is the reference for tumor_region, HPV-positive is the reference for HPV_status, and no chemotherapy is the reference for chemo. GS2 is the gene signature score and GS2_score:HPV_status represents an estimated interaction between the signature score and HPV status.

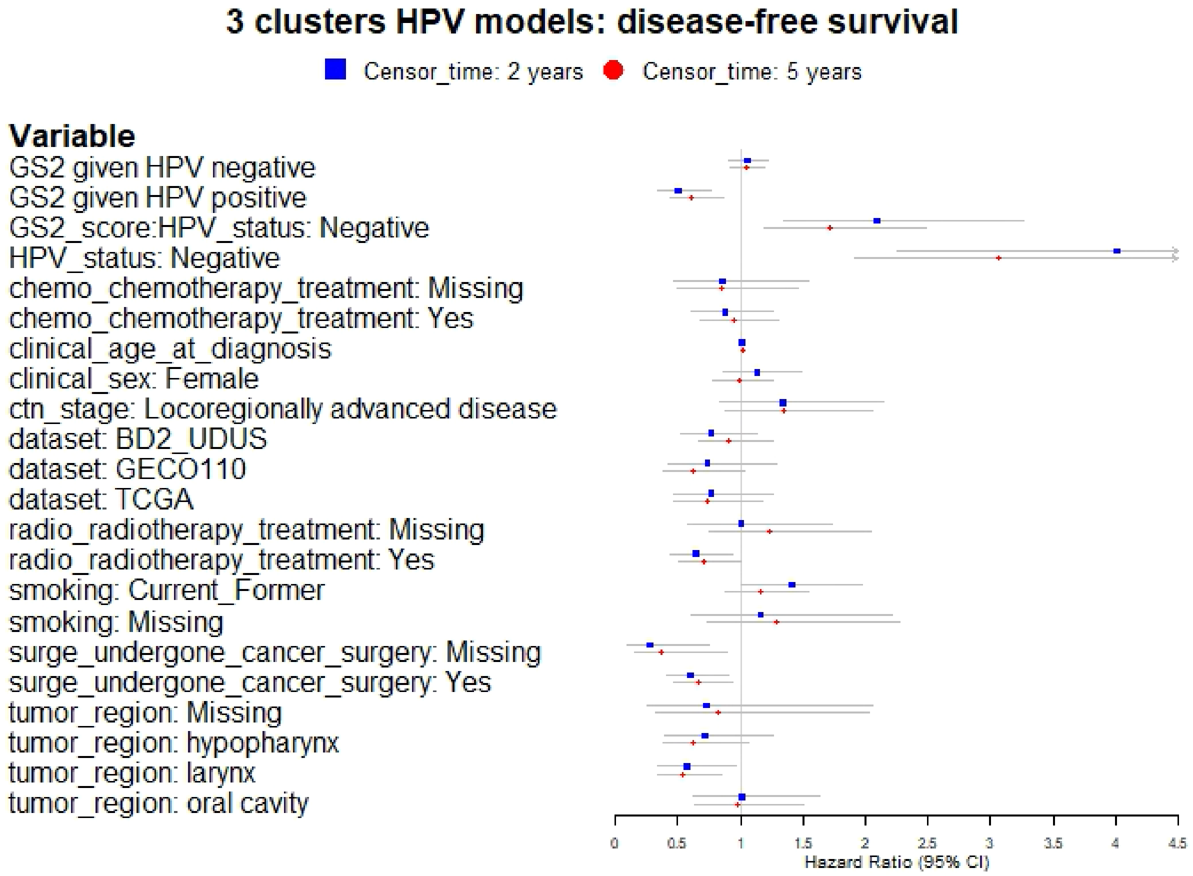

**Figure S7.** Forestplot showing the hazard ratio estimates (with error bars representing 95% confidence intervals) for different variables/parameters in multivariable models testing the *3 clusters HPV* signature where disease-free survival was the endpoint. Two models are compared, where the difference is the censoring time of the endpoint (2 years or 5 years since diagnosis). The arrow on error bars implies that the upper confidence boundary is outside the range of values shown on the plot (see Table S7 for precise estimates). Early disease is the reference for ctn_stage, oropharynx is the reference for tumor_region, HPV-positive is the reference for HPV_status, and no chemotherapy is the reference for chemo. GS2 is the gene signature score and GS2_score:HPV_status represents an estimated interaction between the signature score and HPV status.

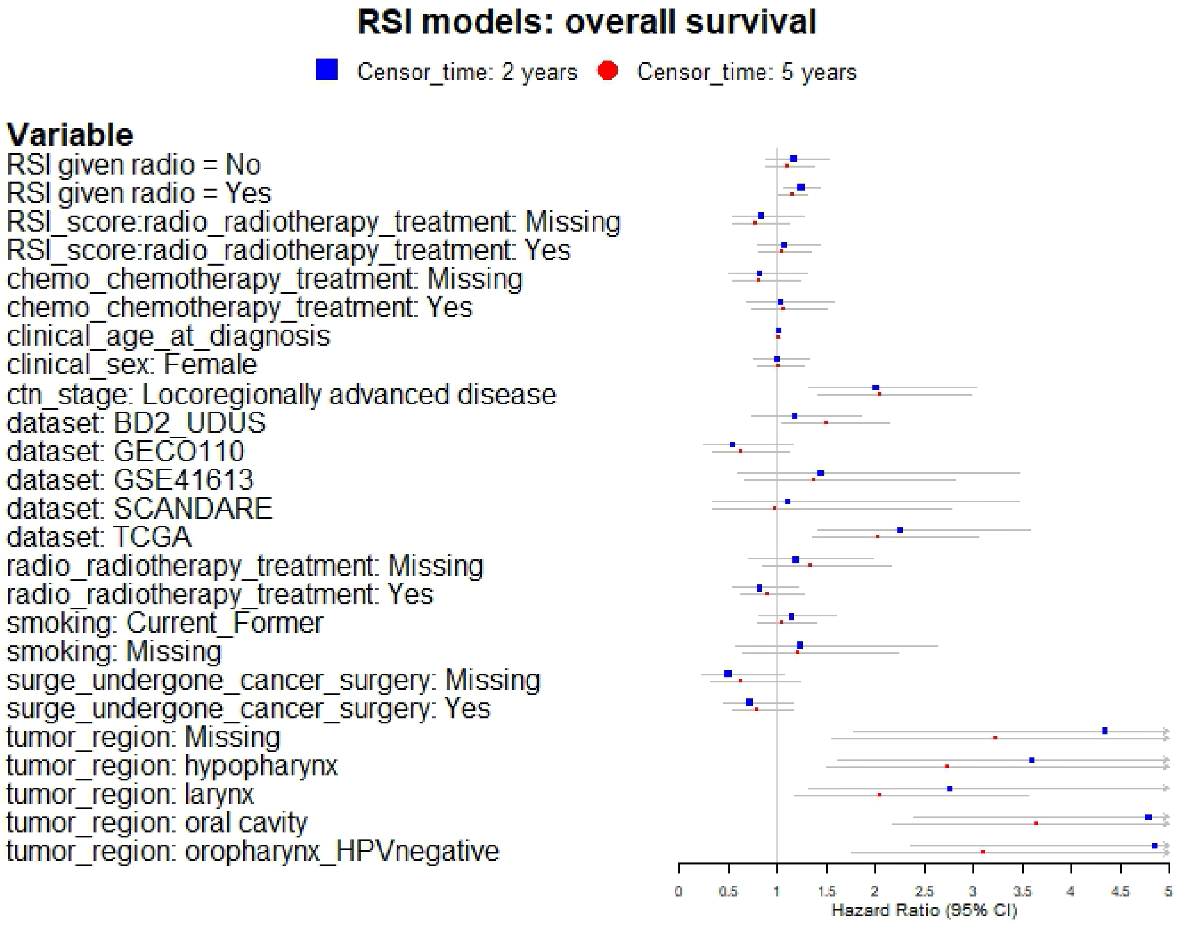

**Figure S8.** Forestplot showing the hazard ratio estimates (with error bars representing 95% confidence intervals) for different variables/parameters in multivariable models testing the radiosensitivity index (*RSI*) where overall survival was the endpoint. Two models are compared, where the difference is the censoring time of the endpoint (2 years or 5 years since diagnosis). The arrow on error bars implies that the upper confidence boundary is outside the range of values shown on the plot (see Table S8 for precise estimates). Early disease is the reference for ctn_stage, HPV-positive oropharynx is the reference for tumor_region, and no chemotherapy is the reference for chemo. RSI is the gene signature score and RSI_score:radio_radiotherapy_treatment represents an estimated interaction between the signature score and radiotherapy where no radiotherapy is the reference.

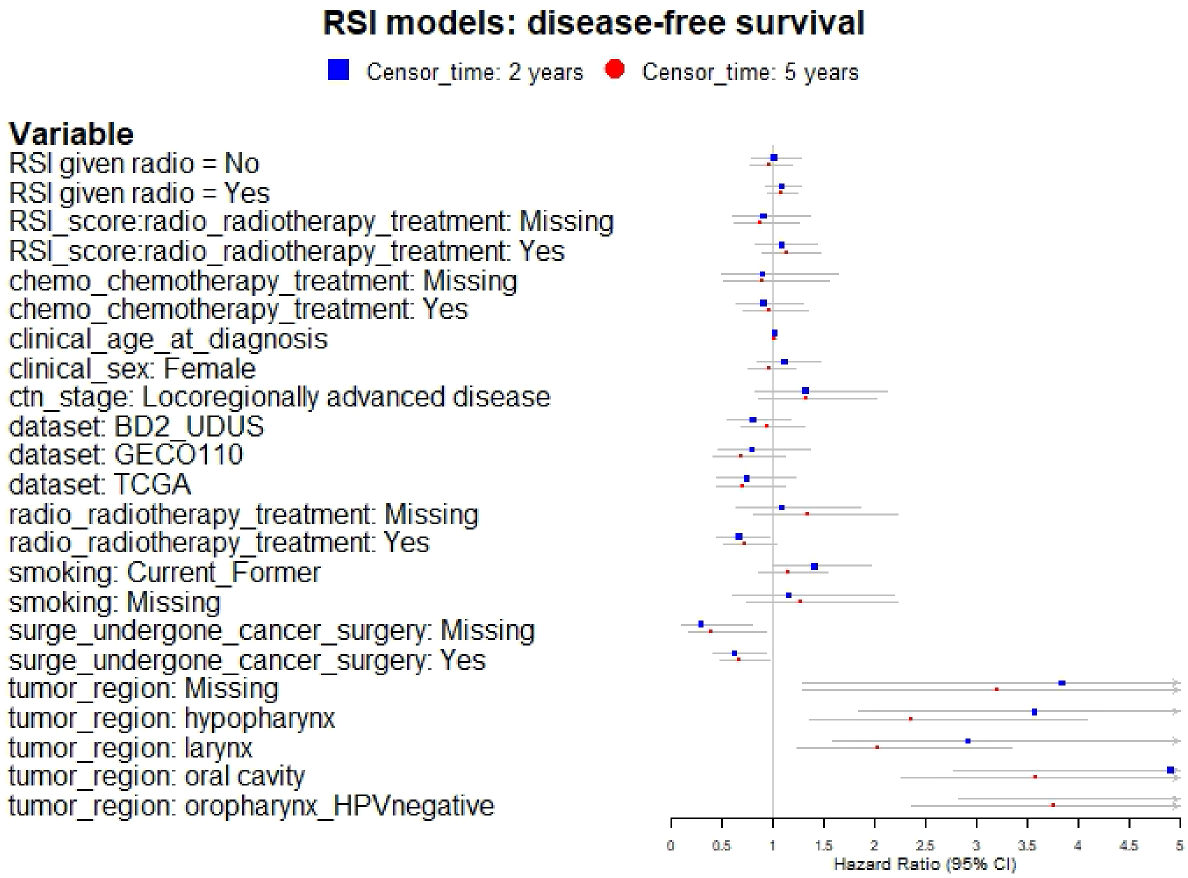

**Figure S9.** Forestplot showing the hazard ratio estimates (with error bars representing 95% confidence intervals) for different variables/parameters in multivariable models testing the radiosensitivity index (*RSI*) where disease-free survival was the endpoint. Two models are compared, where the difference is the censoring time of the endpoint (2 years or 5 years since diagnosis). The arrow on error bars implies that the upper confidence boundary is outside the range of values shown on the plot (see Table S8 for precise estimates). Early disease is the reference for ctn_stage, HPV-positive oropharynx is the reference for tumor_region, and no chemotherapy is the reference for chemo. RSI is the gene signature score and RSI_score:radio_radiotherapy_treatment represents an estimated interaction between the signature score and radiotherapy where no radiotherapy is the reference.

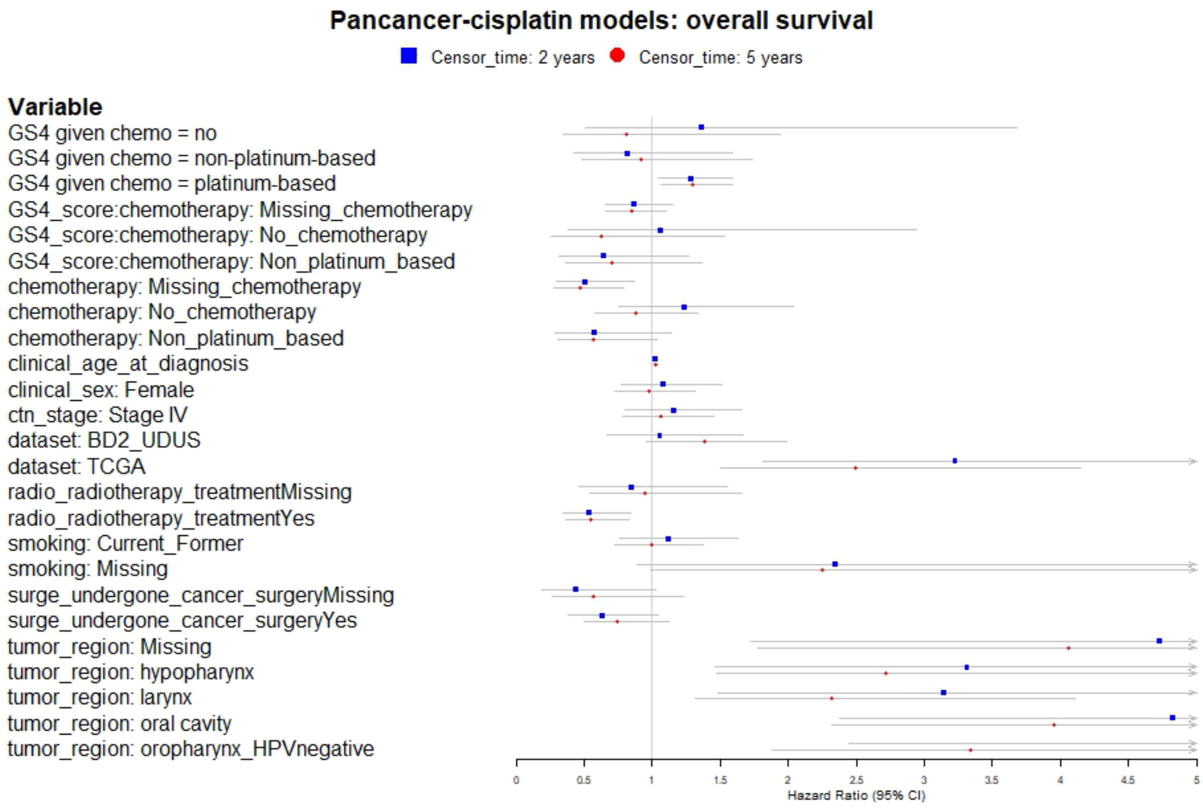

**Figure S10.** Forestplot showing the hazard ratio estimates (with error bars representing 95% confidence intervals) for different variables/parameters in multivariable models testing the *pancancer-cisplatin* signature where overall survival was the endpoint. Two models are compared, where the difference is the censoring time of the endpoint (2 years or 5 years since diagnosis). The arrow on error bars implies that the upper confidence boundary is outside the range of values shown on the plot (see Table S9 for precise estimates). Stage III is the reference for ctn_stage, and HPV-positive oropharynx is the reference for tumor_region. GS4 is the gene signature score and GS4_score:chemotherapy represents an estimated interaction between the signature score and chemotherapy where platinum-based chemotherapy is the reference.

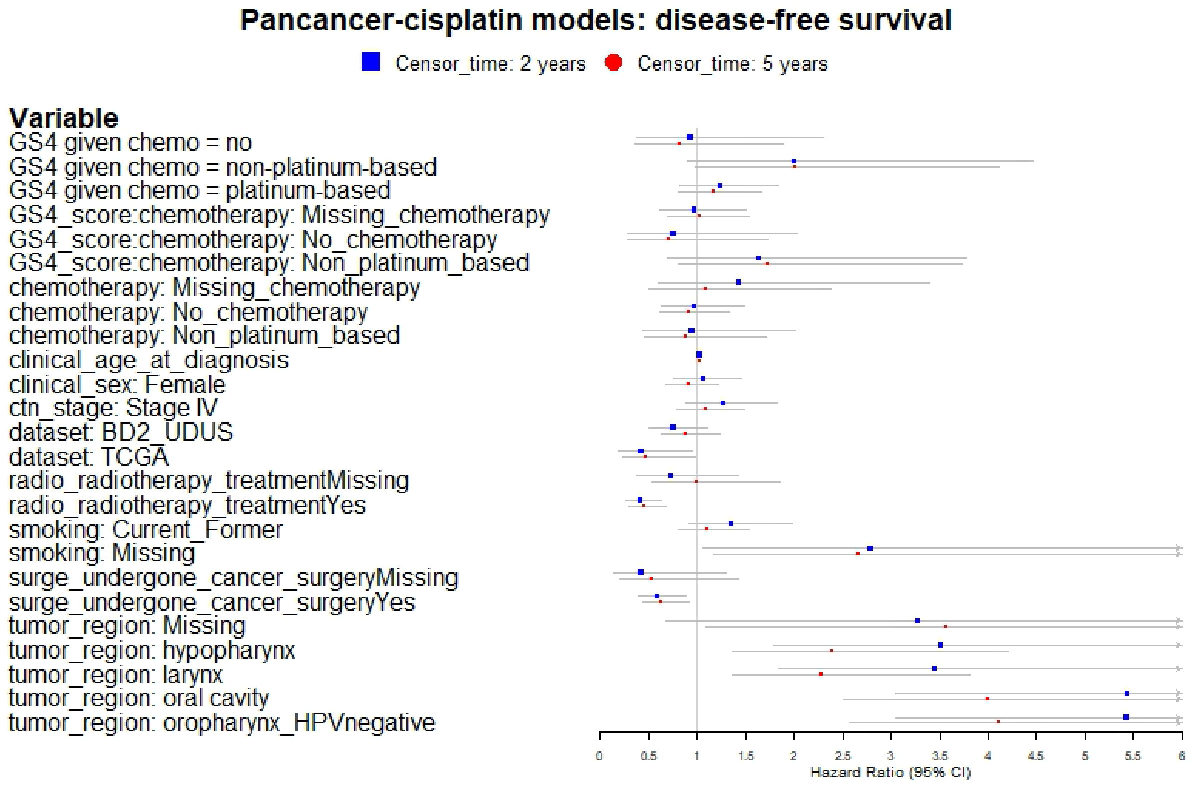

**Figure S11.** Forestplot showing the hazard ratio estimates (with error bars representing 95% confidence intervals) for different variables/parameters in multivariable models testing the *pancancer-cisplatin* signature where disease-free survival was the endpoint. Two models are compared, where the difference is the censoring time of the endpoint (2 years or 5 years since diagnosis). The arrow on error bars implies that the upper confidence boundary is outside the range of values shown on the plot (see Table S9 for precise estimates). Stage III is the reference for ctn_stage, and HPV-positive oropharynx is the reference for tumor_region. GS4 is the gene signature score and GS4_score:chemotherapy represents an estimated interaction between the signature score and chemotherapy where platinum-based chemotherapy is the reference.

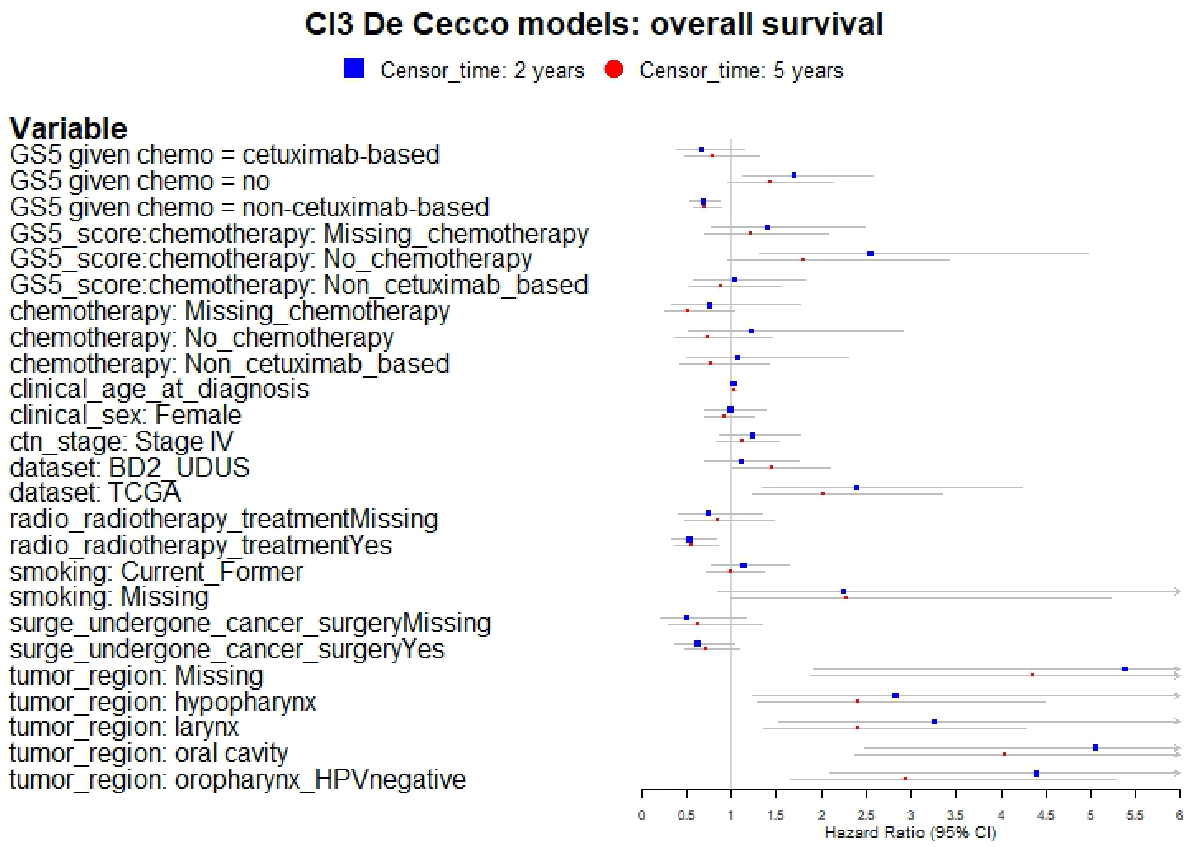

**Figure S12.** Forestplot showing the hazard ratio estimates (with error bars representing 95% confidence intervals) for different variables/parameters in multivariable models testing the *Cl3-hypoxia* signature where overall survival was the endpoint. Two models are compared, where the difference is the censoring time of the endpoint (2 years or 5 years since diagnosis). The arrow on error bars implies that the upper confidence boundary is outside the range of values showed on the plot (see Table S10 for precise estimates). Stage III is the reference for ctn_stage, and HPV-positive oropharynx is the reference for tumor_region. GS5 is the gene signature score and GS5_score:chemotherapy represents an estimated interaction between the signature score and chemotherapy where cetuximab-based treatment is the reference.

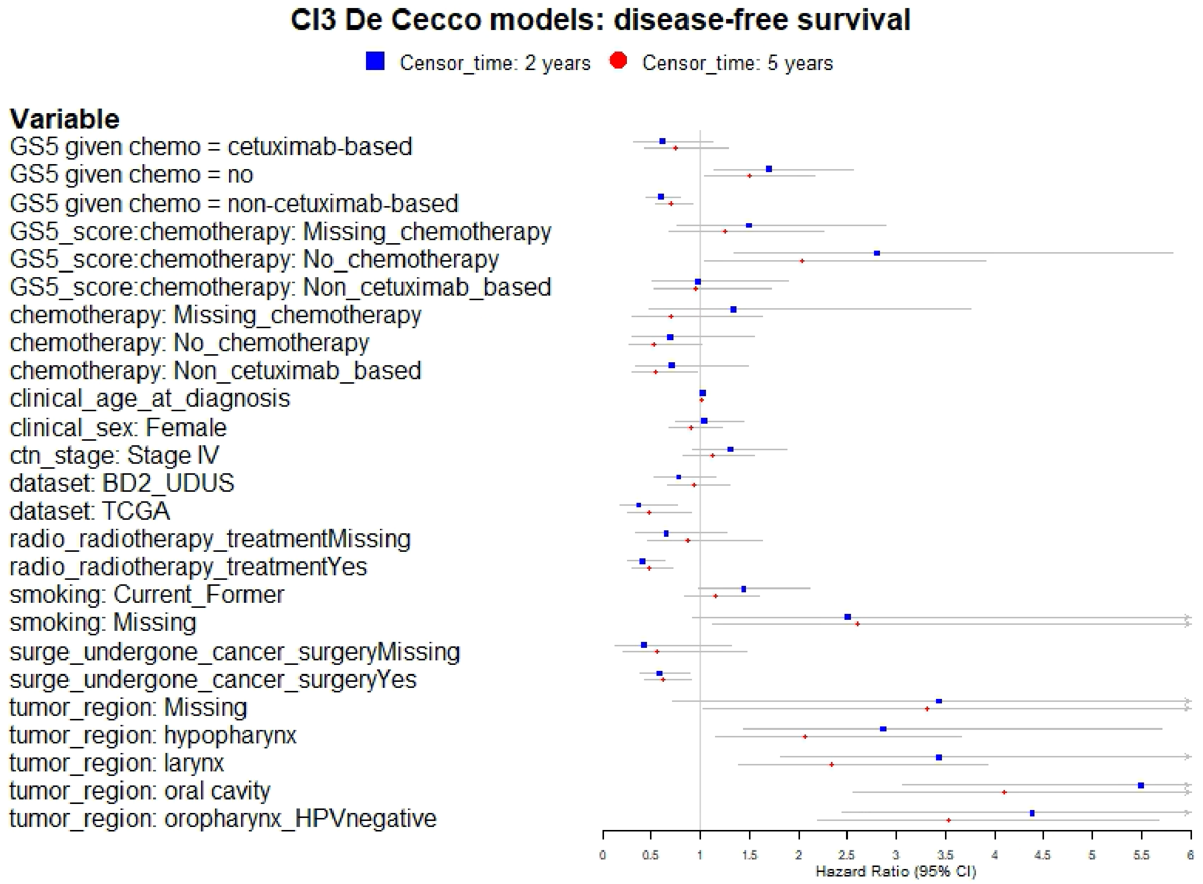

**Figure S13.** Forestplot showing the hazard ratio estimates (with error bars representing 95% confidence intervals) for different variables/parameters in multivariable models testing the *Cl3-hypoxia* signature where disease-free survival was the endpoint. Two models are compared, where the difference is the censoring time of the endpoint (2 years or 5 years since diagnosis). The arrow on error bars implies that the upper confidence boundary is outside the range of values showed on the plot (see Table S10 for precise estimates). Stage III is the reference for ctn_stage, and HPV-positive oropharynx is the reference for tumor_region. GS5 is the gene signature score and GS5_score:chemotherapy represents an estimated interaction between the signature score and chemotherapy where cetuximab-based treatment is the reference.
