## Supplementary Tables S1-S3 for "External validation of prognostic and predictive gene signatures in head and neck cancer patients"

**Table S1.** Datasets screened in this study showing number of screened patients (N) and number of eligible patients per dataset and clinical scenario. Study ID is the ID used in this manuscript. Study short name is original short study name. OS = overall survival. DFS = disease-free survival.

| Study ID | Study short name | Provider | ClinicalTrials.gov ID | N | Eligible for *172-GS, 3 clusters HPV* and *RSI* signatures | | Eligible for *pancancer-cisplatin* and *Cl3-hypoxia* signatures | |
|---|---|---|---|---|---|---|---|---|
| | | | | | OS | DFS | OS | DFS |
| BD2_INT_MI | BD2Decide INT | Istituto Nazionale dei Tumori | NCT02832102 | 310 | 310 | 310 | 310 | 310 |
| BD2_UDUS | BD2Decide UDUS | Charite - subcontractor | NCT02832102 | 127 | 127 | 127 | 127 | 127 |
| GECO110 | GECO110 | Istituto Nazionale dei Tumori | | 107 | 107 | 107 | 0 | 0 |
| Charite_ARO0401 | ARO 04-01 Def-RCT | Charite | NCT02059668 | 58 | 0 | 0 | 0 | 0 |
| Charite_DKTKROdef | DKTKRO Def-RCT | Charite | NCT02059668 | 63 | 0 | 0 | 0 | 0 |
| PROGOR1 | Curie PROGOR 1 | Curie | | 120 | 0 | 0 | 0 | 0 |
| PROGOR3 | Curie PROGOR 3 | Curie | | 99 | 0 | 0 | 0 | 0 |
| SCANDARE | SCANDARE | Curie | NCT03017573 | 80 | 32 | 0 | 0 | 0 |
| TCGA | TCGA-HNSC | Public | | 528 | 424 | 363 | 313 | 263 |
| GSE41613 | GSE41613 | Public | | 97 | 97 | 0 | 0 | 0 |
| Total | SuPerTreat | | | 1589 | 1097 | 907 | 750 | 700 |

**Table S2.** Cohort characteristics for head and neck cancer patients used to test the radiosensitivity index (*RSI*). The disease-free survival (DFS) dataset is a subset of the overall survival dataset, where patients without data on DFS are excluded.

| | Overall survival dataset (N=1097) | Disease-free survival dataset (N=907) |
|---|---|---|
| ***RSI* score** | | |
| Mean (SD) | 0.0 (1.0) | 0.0 (1.0) |
| Median (Min, Max) | -0.1 (-2.5, 4.6) | -0.1 (-2.4, 3.5) |
| **Radiotherapy treatment** | | |
| No | 208 (19.0%) | 146 (16.1%) |
| Yes | 752 (68.6%) | 633 (69.8%) |
| Missing | 137 (12.5%) | 128 (14.1%) |
| **Study ID** | | |
| BD2_INT_MI | 310 (28.3%) | 310 (34.2%) |
| BD2_UDUS | 127 (11.6%) | 127 (14.0%) |
| GECO110 | 107 (9.8%) | 107 (11.8%) |
| GSE41613 | 97 (8.8%) | 0 (0%) |
| SCANDARE | 32 (2.9%) | 0 (0%) |
| TCGA | 424 (38.7%) | 363 (40.0%) |
| **Clinical age at diagnosis** | | |
| Mean (SD) | 61.0 (11.9) | 60.8 (11.7) |
| Median (Min, Max) | 61.0 (19.0, 93.0) | 61.0 (19.0, 93.0) |
| **Clinical sex** | | |
| Male | 794 (72.4%) | 660 (72.8%) |
| Female | 303 (27.6%) | 247 (27.2%) |
| **cTN disease extension at diagnosis** | | |
| Early disease | 161 (14.7%) | 98 (10.8%) |
| Locoregionally advanced disease | 936 (85.3%) | 809 (89.2%) |
| **Tumor region and HPV-status** | | |
| Oropharynx and HPV-positive | 156 (14.2%) | 151 (16.6%) |
| Hypopharynx | 55 (5.0%) | 52 (5.7%) |
| Larynx | 165 (15.0%) | 150 (16.5%) |
| Oral cavity | 598 (54.5%) | 451 (49.7%) |
| Oropharynx and HPV-negative | 71 (6.5%) | 67 (7.4%) |
| Missing | 52 (4.7%) | 36 (4.0%) |
| **Smoking** | | |
| Never | 230 (21.0%) | 213 (23.5%) |
| Current_Former | 653 (59.5%) | 609 (67.1%) |
| Missing | 214 (19.5%) | 85 (9.4%) |
| **Undergone cancer surgery** | | |
| No | 187 (17.0%) | 187 (20.6%) |
| Yes | 854 (77.8%) | 672 (74.1%) |
| Missing | 56 (5.1%) | 48 (5.3%) |
| **Systemic treatment** | | |
| No | 302 (27.5%) | 233 (25.7%) |
| Yes | 461 (42.0%) | 426 (47.0%) |
| Missing | 334 (30.4%) | 248 (27.3%) |

**Table S3.** Cohort characteristics for head and neck cancer patients used to test gene signatures *pancancer-cisplatin* and *Cl3-hypoxia*. The disease-free survival (DFS) dataset is a subset of the overall survival dataset, where patients without data on DFS are excluded.

| | Overall survival dataset (N=750) | Disease-free survival dataset (N=700) |
|---|---|---|
| ***Pancancer-cisplatin* score** | | |
| Mean (SD) | 0.0 (1.0) | 0.0 (1.0) |
| Median (Min, Max) | 0.0 (-4.8, 4.8) | 0.0 (-5.1, 4.8) |
| ***Cl3-hypoxia* score** | | |
| Mean (SD) | 0.0 (1.0) | 0.0 (1.0) |
| Median (Min, Max) | 0.0 (-3.2, 3.1) | 0.0 (-3.4, 3.2) |
| **Systemic agent (platinum)** | | |
| Platinum-based | 362 (48.3%) | 338 (48.3%) |
| Non-platinum-based | 51 (6.8%) | 47 (6.7%) |
| No systemic treatment | 155 (20.7%) | 154 (22.0%) |
| Missing systemic treatment | 182 (24.3%) | 161 (23.0%) |
| **Systemic agent (cetuximab)** | | |
| Cetuximab-based | 35 (4.7%) | 32 (4.6%) |
| Non-cetuximab-based | 378 (50.4%) | 353 (50.4%) |
| No systemic treatment | 155 (20.7%) | 154 (22.0%) |
| Missing systemic treatment | 182 (24.3%) | 161 (23.0%) |
| **Study ID** | | |
| BD2_INT_MI | 310 (41.3%) | 310 (44.3%) |
| BD2_UDUS | 127 (16.9%) | 127 (18.1%) |
| TCGA | 313 (41.7%) | 263 (37.6%) |
| **Clinical age at diagnosis** | | |
| Mean (SD) | 61.2 (11.0) | 61.2 (11.1) |
| Median (Min, Max) | 61.0 (20.0, 93.0) | 61.0 (20.0, 93.0) |
| **Clinical sex** | | |
| Male | 560 (74.7%) | 522 (74.6%) |
| Female | 190 (25.3%) | 178 (25.4%) |
| **Tumor stage (TNM 7th edition)** | | |
| Stage III | 177 (23.6%) | 167 (23.9%) |
| Stage IV | 573 (76.4%) | 533 (76.1%) |
| **Tumor region and HPV-status** | | |
| Oropharynx and HPV-positive | 145 (19.3%) | 144 (20.6%) |
| Hypopharynx | 53 (7.1%) | 51 (7.3%) |
| Larynx | 149 (19.9%) | 139 (19.9%) |
| Oral cavity | 303 (40.4%) | 274 (39.1%) |
| Oropharynx and HPV-negative | 65 (8.7%) | 65 (9.3%) |
| Missing | 35 (4.7%) | 27 (3.9%) |
| **Smoking** | | |
| Never | 188 (25.1%) | 173 (24.7%) |
| Current or Former | 550 (73.3%) | 515 (73.6%) |
| Missing | 12 (1.6%) | 12 (1.7%) |
| **Undergone cancer surgery** | | |
| No | 187 (24.9%) | 187 (26.7%) |
| Yes | 519 (69.2%) | 475 (67.9%) |
| Missing | 44 (5.9%) | 38 (5.4%) |
| **Radiotherapy treatment** | | |
| No | 84 (11.2%) | 79 (11.3%) |
| Yes | 585 (78.0%) | 545 (77.9%) |
| Missing | 81 (10.8%) | 76 (10.9%) |
